# Ultra-rare biallelic *THAP12* variants cause loss of function and underlie severe epileptic encephalopathy

**DOI:** 10.64898/2026.02.27.26347078

**Authors:** Katarzyna Ochenkowska, Bryce Rampal, Antoine Légaré, Valérie Triassi, Sébastien Audet, Alexandre Brisson, Guillaume Bernas, Meijiang Liao, Alexandra da Silva Babinet, Julie Pilliod, Mariah Gillaspie, Grace E. VanNoy, Lynn Pais, Anne O’Donnell-Luria, Nicole Leclerc, Diana Walleigh, Jean-François Schmouth, Laurent Cappadocia, Patrick Desrosiers, Paul De Koninck, Martine Tétreault, Éric Samarut

**Affiliations:** Department of Neuroscience, Faculty of Medicine, Université de Montréal, Montreal, H3T 1J4, Quebec, Canada; University of Montréal Hospital Research Center (CRCHUM), Montreal H2X 0A9, Quebec Canada; CERVO Brain Research Centre, Université Laval, Québec, G1J 2G3, Canada; Université du Québec à Montréal, Montréal, QC, Canada; Quebec Network for Research on Protein Function, Engineering, and Applications (PROTEO), Montréal, QC, Canada; The Lightning and Love Foundation; Center for Mendelian Genomics, Program in Medical and Population Genetics, Broad Institute of MIT and Harvard, Cambridge, MA, USA; Division of Genetics and Genomics, Boston Children’s Hospital, Harvard Medical School, Boston, MA, USA; Center for Genomic Medicine, Massachusetts General Hospital, Harvard Medical School, Boston, MA, USA; Section of Child Neurology, Department of Pediatrics, University of Colorado, Aurora, CO, USA; Centre d’excellence en recherche sur les maladies orphelines – Fondation Courtois (CERMO-FC), Université du Québec à Montréal, Montreal H2X 3Y7, Quebec Canada

**Keywords:** Developmental Epileptic Encephalopathies, Infantile Spasms (West syndrome), Lennox-Gastaut Syndrome, Functional Genomics, THAP12, Neurodevelopment

## Abstract

Developmental and epileptic encephalopathies (DEEs) are a group of severe childhood-onset neurological disorders, often caused by rare genetic variants affecting brain development and excitability. Despite advances in genomic sequencing, a substantial proportion of DEE cases remain unsolved. Here, we identify *THAP12* as a novel disease-causing gene associated with autosomal recessive DEE. Whole-genome sequencing in two siblings who presented with infantile spasms and progressed to Lennox-Gastaut syndrome revealed compound heterozygous variants in *THAP12*, leading to a reduction in protein abundance, consistent with a loss-of-function mechanism. To confirm this mechanism *in vivo*, we generated mouse models carrying either of the two patient-specific alleles. Both homozygous and compound heterozygous animals exhibited embryonic lethality, confirming the essential and dosage-sensitive role of *Thap12* during early development. Zebrafish loss-of-function models recapitulated major aspects of the human phenotype, including microcephaly, brain hypoplasia, abnormal neuronal activity, and increased seizure sensitivity. Transcriptomic profiling of larval zebrafish brains revealed dysregulation of cell cycle and apoptotic pathways, in line with increased cell death and reduced proliferation observed in mutant embryos. Notably, overexpression of wild-type human *THAP12* mRNA rescued these *in vivo* phenotypes, while the patient-derived variants allele failed to do so. Altogether, our findings demonstrate that *THAP12* is essential for early brain development and neuronal survival, and that biallelic loss-of-function variants in this gene underlie a previously unrecognized etiology of autosomal recessive DEE. These results provide a mechanistic framework linking, for the first time, *THAP12* dysfunction to neurodevelopmental pathology and open new avenues for diagnosis in undiagnosed DEE cases.

## Introduction

Developmental and epileptic encephalopathies (DEEs) are severe pediatric neurological conditions, presenting early in life with drug-resistant seizures and varying degrees of developmental delay or cognitive impairment^1-3^. Genetic causes are increasingly recognized as major contributors to these disorders^4-6^. Identifying the underlying genetic variant can guide medical management, improve prognostic accuracy, and support genetic counselling^7,8^. However, despite the growing use of gene panels and next-generation sequencing in clinical practice, at least a quarter of DEE patients remain without a molecular diagnosis^9-11^. Among these conditions, infantile spasms with progression to Lennox-Gastaut syndrome represent particularly severe and complex forms^12-14^. Infantile spasms typically begin within the first year of life and may evolve into Lennox-Gastaut syndrome^15^, which is characterized by multiple seizure types, intellectual disability, and distinct electroencephalogram (EEG) patterns including slow spike and wave with paroxysmal fast activity during sleep^16^. These syndromes carry a high medical burden^2^ and are often refractory to treatment.

In this context, uncovering novel genes and variants involved in these disorders remains a major challenge and priority. Indeed, in addition to improving the diagnostic capabilities available to undiagnosed patients, this opens the door to a better understanding of the underlying pathogenic mechanisms. This is especially important in the case of epileptic seizures that are resistant to currently available treatments, suggesting that new molecular targets should be explored for the development of alternative therapies. Importantly, conventional diagnostic panels remain targeted and can therefore miss ultra-rare or novel gene variations. Studying families with multiple affected children can help identify new candidate genes, particularly under recessive inheritance models. However, the lack of recurrence of these ultra-rare variants across multiple families could make their pathogenic interpretation difficult and require multiple levels of functional validation in complementary system models.

In this study, we report a non-consanguineous family with two siblings affected by a severe developmental epileptic encephalopathy presenting with infantile spasms and progressing to the Lennox-Gastaut continuum^17^. Despite inconclusive genetic panels, the occurrence of the same clinical presentation in two sisters from healthy parents is consistent with and suggestive of a recessive genetic origin. We therefore performed whole-genome sequencing (WGS) and identified compound heterozygous variants in *THAP12*, a gene not previously associated with neurological disease. *THAP12* is a vertebrate-only gene and has been reported to have low tissue specificity in humans, including in the brain (Human Protein Atlas, The Genotype-Tissue Expression Portal), suggesting broad, ubiquitous expression. It is a member of the Thanatos Associated Protein family, a group involved in several key cellular functions, such as transcription regulation^18-20^, cell cycle regulation^21-24^, apoptosis^25-28^, and tumor formation^29-31^. Recently, *THAP12* has been identified as a molecular driver of lymphoproliferation^32^, but to date, it has never been associated with any neurological diseases or neurodevelopmental pathways. Despite this, the Whole Genome Sequencing (WGS) analysis of these two affected sisters converges on this gene, and we hypothesize that it may represent a yet undiscovered candidate gene associated with DEE. To investigate the pathogenicity of these variants, we performed a comprehensive series of functional studies across multiple complementary biological systems. These included *in silico* structural modeling, transcriptomic and protein-level assays in patient-derived primary fibroblasts, genetically humanized mouse models carrying patient-specific *THAP12* variants and CRISPR-based zebrafish genetic models. Our results show that patients’ variants result in a drastic reduction in THAP12 protein abundance, suggesting a loss-of-function mechanism consistent with the recessive inheritance pattern. Across all *in vivo* models, we showed that loss of *THAP12* function led to consistent and convergent neurodevelopmental phenotypes, including premature lethality, microcephaly, abnormal neural activity, and increased seizure susceptibility. Importantly, these phenotypes could be rescued by WT *THAP12* cDNA expression, but not by *THAP12* cDNA carrying our patients’ variants.

Altogether, our findings support *THAP12* as a novel disease gene involved in early brain development and whose loss-of-function is causing early-onset epileptic encephalopathy. Beyond this clinical implication, our work also positions *THAP12* as a key regulator of neurogenesis, laying the foundation for future studies into its biological function and its broader relevance to brain disorders.

## Results

### Unresolved genetic diagnosis in two siblings with Infantile Spasms and Lennox-Gastaut syndrome

We describe two female siblings born to healthy, non-consanguineous parents (Fig. 1A). Both pregnancies and deliveries were uneventful. The first sibling (Proband II-1) required an eight-day stay in the neonatal intensive care unit for feeding difficulties, whereas the second sibling (Proband II-2) was discharged home without complications. Early development in the first weeks of life appeared normal. Both children presented with epileptic spasms during early infancy (before 12 months of age). EEGs revealed hypsarrhythmia in both cases (Fig. 1B, 1C), consistent with a diagnosis of infantile spasms. The epilepsy proved refractory to medical treatment in both siblings and evolved into a phenotype consistent with Lennox-Gastaut syndrome (LGS). EEG findings during follow-up were diffusely slow, poorly organized, and featured generalized slow spike-and-wave activity and paroxysmal fast activity during sleep meeting criteria for LGS (Fig. 1D, 1E). Seizures remained poorly controlled despite multiple antiseizure medications. Proband II-1 was treated with levetiracetam, Adrenocorticotropic Hormone (ACTH), prednisone, vigabatrin, clobazam, topiramate, cannabidiol, lamotrigine, lacosamide, and a ketogenic diet. Proband II-2 received ACTH, pulse dexamethasone, clobazam, cannabidiol, levetiracetam, vigabatrin, and lacosamide. Following three treatment courses with ACTH, monthly pulse corticosteroid therapy was initiated to suppress relapses. Seizure frequency in both sisters ranged from daily to weekly, with worsening during intercurrent illness, stress, or fatigue. In parallel with the epileptic manifestations, both children exhibited profound neurodevelopmental delay. They remain nonverbal and non-ambulatory, with complete dependence for all activities of daily living. Neither proband achieved independent mobility, and purposeful use of limbs is minimal. Proband II-1 shows almost no voluntary movement. Muscle tone is markedly abnormal, with a mixed pattern of hypertonia and hypotonia. Hypertonic areas are treated with oral baclofen.

**Figure 1.**
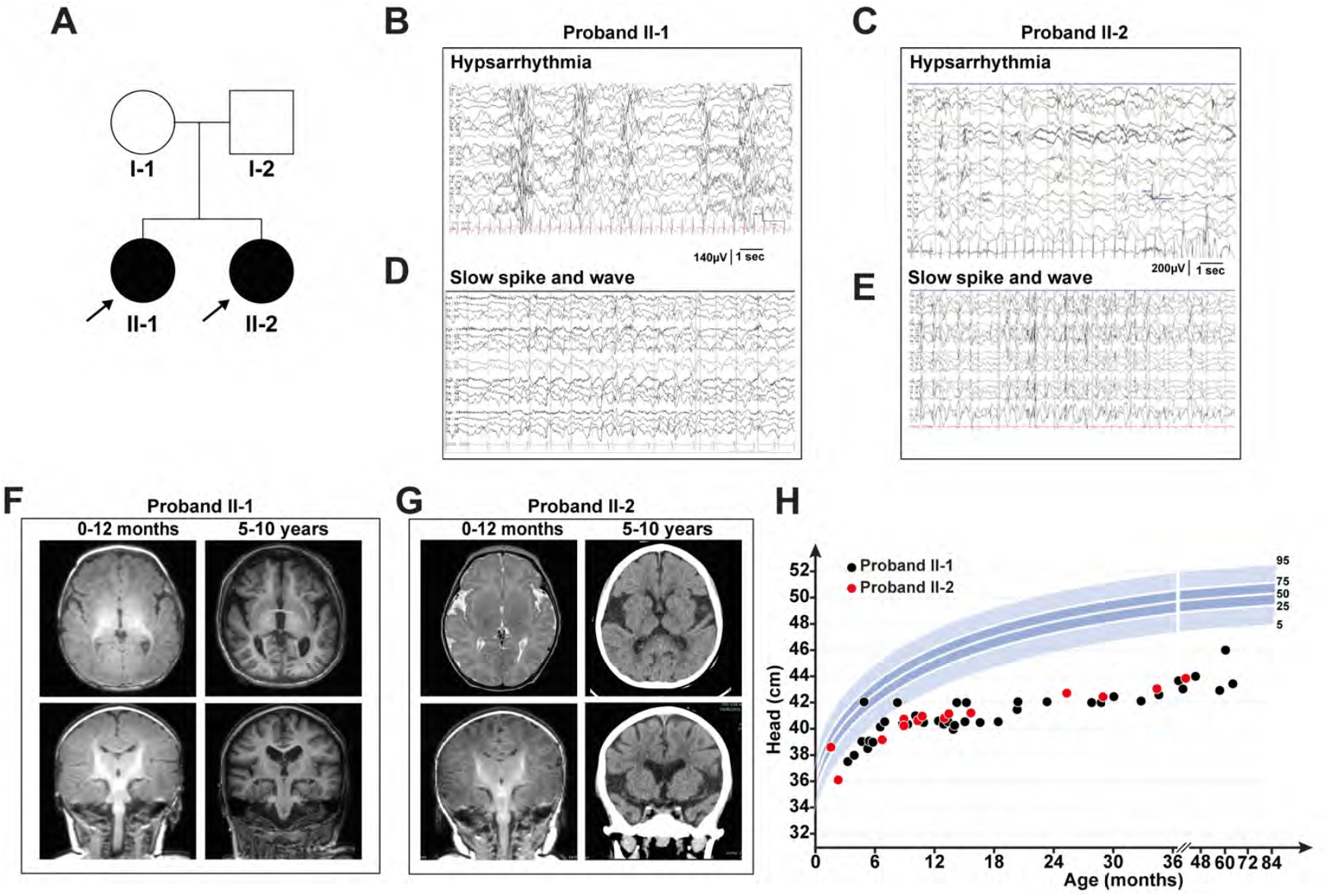
Clinical overview of two siblings with infantile spasms and progression to Lennox-Gastaut syndrome. **(A)** Pedigree of the family showing two affected sisters born to healthy, non-consanguineous parents. **(B-C)** EEG recordings during infancy (0-12 months) reveal hypsarrhythmia in both probands, consistent with a diagnosis of infantile spasms. **(D-E)** Subsequent EEGs in early childhood (1-5 years) show generalized slow spike-and-wave discharges and paroxysmal fast activity during sleep, consistent with Lennox-Gastaut syndrome. **(F-G)** Brain MRI and CT scans in early infancy (0-12 months) and late childhood (5-10 years) reveal moderate cerebral and cerebellar volume loss in both probands. **(I)** Head circumference measurements plotted over time confirm postnatal microcephaly in both siblings, with values below the 2^nd^ percentile (percentiles shown in blue).

Brain MRI confirmed microcephaly without overt malformations or etiologic structural anomalies. In Proband II-1, imaging in laye childhood (5-10 years of age) revealed moderate cerebral and cerebellar volume loss compared to imaging in early infancy (0-12 months) with small hippocampi and symmetric loss of internal architecture (Fig. 1F). In Proband II-2, MRI in early infancy was followed by a CT scan in late childhood, which revealed diffuse parenchymal volume loss with marked cerebellar and brainstem atrophy compared to the earlier imaging (Fig. 1G). Neurological examination revealed generalized hypotonia and poor head control. Progressive microcephaly was noted in both siblings, with serial head circumference measurements consistently below the 2nd percentile (Fig. 1H). Additional clinical features included cortical visual impairment, feeding difficulties requiring gastrostomy tube placement, and frequent respiratory complications, including poor secretion clearance. Cortical visual impairment is present in both sisters, with no consistent visual tracking or fixation, though brief responses to familiar or high-contrast stimuli are occasionally noted. Both siblings required nocturnal ventilatory support via average volume-assured pressure support (AVAPS) to promote lung expansion and oxygenation. Supplemental oxygen is administered intermittently or continuously based on clinical needs. They experience chronic respiratory disease characterized by hypoventilation, recurrent infections, and difficulty maintaining lung expansion. Recurrent respiratory illnesses often require hospitalization, pressure escalation, and sometimes intubation. Airway clearance involves chest physiotherapy, suctioning, and nebulized treatments multiple times daily. Diagnostic workup included metabolic screening, chromosomal microarray analysis, and epilepsy gene panels targeting known DEE genes. All clinical investigations were non-diagnostic. However, the recurrence of an almost identical severe phenotype in two siblings born to unaffected parents strongly suggested a monogenic, likely recessive etiology.

### Family-based whole-genome sequencing identified rare recessive variants in THAP12

To identify the underlying genetic cause, we performed WGS on whole blood derived from both affected siblings and their unaffected parents. Variant filtering was conducted under both a recessive and dominant inheritance model, prioritizing variants shared by both siblings and for the recessive model, inherited from each heterozygous parent (in trans). Following iterative filtering and prioritization, the final candidate variants reported in this study were absent from population databases (absent from gnomAD v2.1.1 and extremely rare in gnomAD v4.1.0, p.Pro277Thr present in 2 individuals with European ancestry in v4.1.0). Variant prioritization was primarily guided by inheritance model, predicted functional consequence, gene relevance, and consistency with the clinical phenotype. *In silico* prediction scores were used as supportive annotations to aid interpretation rather than as strict filtering criteria, and thresholds were considered within a research-based exploratory framework rather than for clinical diagnostic interpretation. (GERP > 4, PhastCons > 400, SIFT > 0.5, Polyphen-2 > 0.95, CADD>15)^33^. From this list, we identified a single gene, *THAP12* (ENSG00000137492, NM_004705.2 aka *THAP0, DAP4, PRKRIR p52rIPK*), which harbored two variants shared by both sisters, inherited independently from each parent: a paternally-inherited missense variant (c.829C>A / p.Pro277Thr) and a maternally-inherited frameshift deletion (c.313GA>G / p.Glu105AsnfsTer2) (Fig. S1B, Fig. 2A). We entered the *THAP12* gene and sibling’s phenotypes into Matchmaker Exchange^34^ through the seqr platform^35^ but were unable to identify any additional patients with biallelic variation in this gene. We also reached out to a number of research teams working on gene discovery to check their datasets without success of identifying additional patients. Allelic ratio analyses from WGS and RNA-seq data (Fig. S1B, S1C) confirmed compound heterozygosity in both patients. While the missense allele was expressed at near-balanced levels, the frameshift allele showed marked transcript depletion, consistent with nonsense-mediated decay. Although these variants are absent from ClinVar and rare in gnomAD, selected *in silico* prediction tools classified the missense variant c.829C>A (p.Pro277Thr) as deleterious (MutationTaster: disease-causing; AlphaMissense score: 0.7272). Moreover, Pro277 is extremely conserved among vertebrates (Fig. 2B), suggesting that it has been under strong selective pressure during evolution due to its important role within the THAP12 protein.

**Figure 2.**
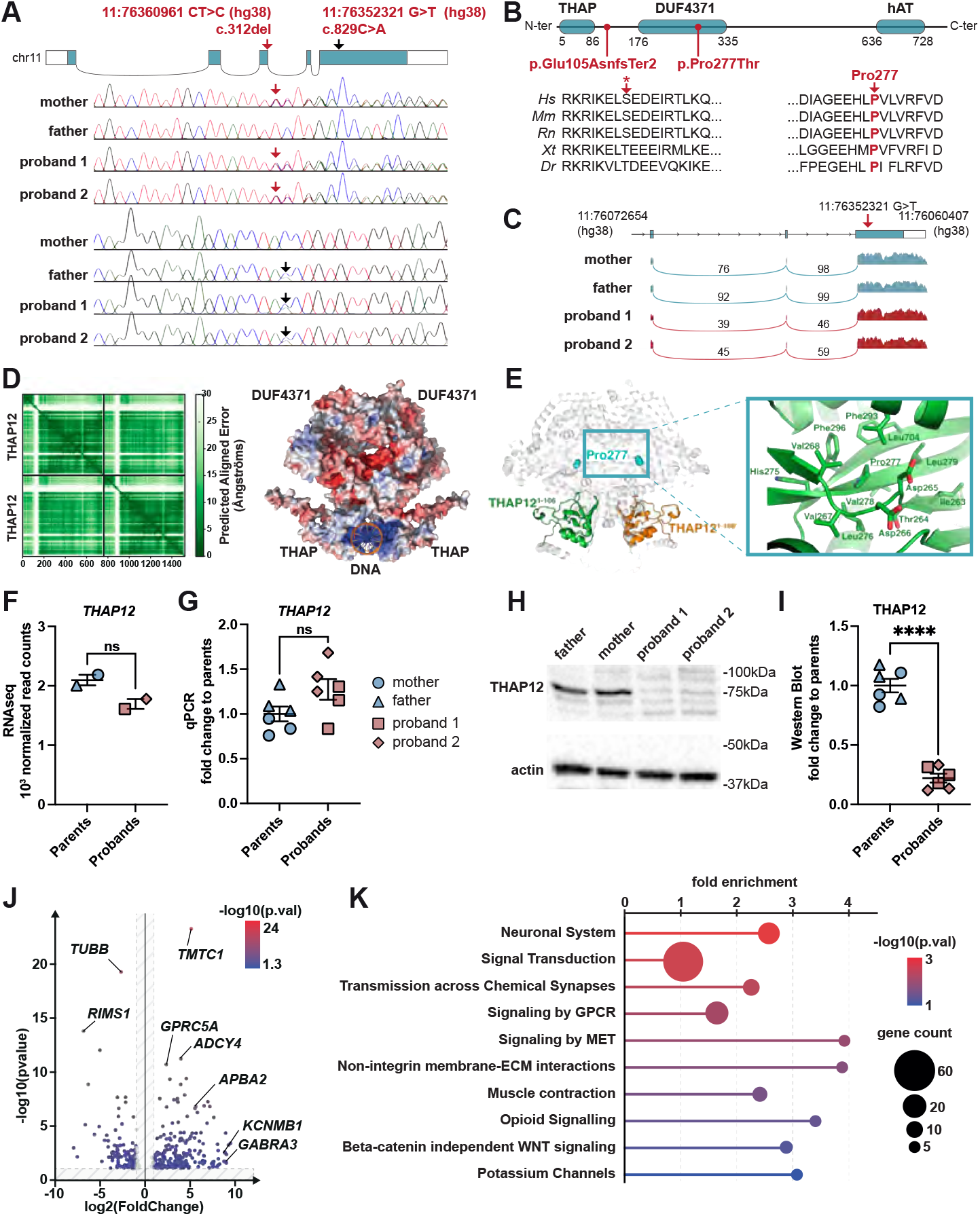
*THAP12* compound heterozygous variants identified in two siblings impair protein stability and impact normal gene expression. (A) Sanger sequencing confirms compound heterozygous variants in *THAP12* in both affected siblings, with a maternally inherited frameshift (c.312del, red arrow) and a paternally inherited missense variant (c.829C>A, black arrow). (B) The two variants affect conserved residues within protein domains, particularly a proline at position 277 in the DUF4371 domain, as shown in a multi-species alignment (red arrow; *Hs: Homo sapiens; Mm: Mus musculus; Rn: Rattus norvegicus; Xt: Xenopus tropicalis; Dr: Danio rerio*). (C) A Sashimi plot of RNA-seq reads from patients’ primary fibroblasts across the *THAP12* locus shows no major changes in exon usage or alternative splicing between probands and their parents. Read counts on the splice junction arcs indicate the number of split reads supporting each exon-exon connection in each sample. **(D)** Structural modelling with AlphaFold3 predicts that THAP12 forms homodimers primarily through interactions between DUF4371 domains (left). In a THAP12-DNA complex predicted using AlphaFold3, the N-terminal THAP zinc-finger domains interacts with DNA using an electropostive surface (right). **(E)** The maternally inherited frameshift variant (Glu105AsnfsTer2) is predicted to truncate the protein after residue 106, abolishing the DUF4371 domain and likely impairing dimerization. The N-terminal segment (residues 1-106) is shown in color, corresponding to the truncated product of the frameshift allele. The paternally inherited Pro277Thr missense variant affects a conserved residue buried within the hydrophobic core of the DUF4371 domain, likely disrupting local folding and protein stability. **(F-G)** *THAP12* transcript levels are not significantly changed in patient fibroblasts compared to parental controls, as shown by RNA-seq and qPCR analyses. **(H-I)** In contrast, THAP12 protein levels in patient fibroblasts are significantly reduced in both probands, as shown by Western blot and quantification. **(J)** Volcano plot displaying differentially expressed genes from bulk RNA-sequencing analysis of patient-derived fibroblasts compared to parental controls. Downregulated genes include several involved in neuronal and synaptic function (e.g., *TUBB, RIMS1, GABRA3, KCNMB1*). Significance is color-coded according to the - log10(adjusted p-value). The full list of differentially expressed genes is provided in Table S1. **(K)** Pathway enrichment analysis of differentially expressed genes highlights over-representation of pathways such as “Neuronal System”, “Signal Transduction”, and “Transmission across Chemical Synapses”. Dot size indicates the number of genes in each pathway; color represents -log10(p-value). *Statistical analyses in panels F, G, and I used unpaired two-tailed Student’s t-test: *p < 0*.*05, **p < 0*.*01, ***p < 0*.*001, ****p < 0*.*0001; ns, not significant*.

### Structural and expression analyses reveal functional impact of patient THAP12 variants

*THAP12* is a ubiquitously expressed, vertebrate-specific gene whose role in neurodevelopment remains unexplored. It encodes a 761-amino-acid-long protein that encompasses three main functional domains: a C2CH zinc finger THAP domain, a Domain of Unknown Function (DUF) 4371, and a Hungerford-Hathaway-Transposase (hAT) dimerization domain. The c.313GA>G identified in our patients results in a change in the protein sequence starting from position 105, causing a premature stop at position 107 (p.Glu105AsnfsTer2; Fig. 2B). The c.829C>A variant leads to the substitution of proline at position 277 with threonine (Pro277Thr, Fig. 2B). Moreover, the c.829C>A variant could introduce a cryptic splice site within exon 5, potentially altering the normal splicing pattern of *THAP12* transcripts. We conducted RNA sequencing using patient-derived primary fibroblasts; however, no alternative splicing was observed in our RNA-seq data analysis (Fig. 2C). We then initiated structural predictions of THAP12 using AlphaFold3^36^ to investigate the molecular impacts of the Pro277Thr and Glu105AsnfsTer2 mutations. Consistently with previous findings^25^, the Predicted Aligned Error (PAE) plot suggests that THAP12 likely forms homodimers (Fig. 2D). Dimerization primarily occurs through interactions between adjacent DUF domains while the N-terminal zinc-binding domains (THAP) fold back on the DUF domain (Fig. S2A). As the THAP domain of other THAP proteins was shown to bind DNA^18,37^, we also modeled the THAP12 dimer with DNA using AlphaFold3. As the exact sequences bound by THAP12 are still elusive, we used a random DNA sequence and obtained a model where a change in orientation of the THAP domain (compare Fig. S2A and S2B) reveals an electropositive surface that engages DNA (Fig. 2D). This mode of DNA binding is generally similar to that of the THAP domain from *Drosophila melanogaster* P-element transposase (pdb 3KDE). The Glu105AsnfsTer2 mutation is located in the linker connecting the N-terminal zinc-binding and DUF domains, likely resulting in a 106 amino-acid long protein lacking the entire DUF domain, which might impede dimerization and prevent normal THAP12’s functions (Fig. 2E). The Pro277Thr variant is found within the DUF domain (Fig. 2B and 2E). In human THAP12, Pro277 is situated in a hydrophobic core of the DUF domain, surrounded by the side chains of Val268, Phe293, Phe296, and Leu704, along with the aliphatic portion of Asp266 (Fig. 2E). This core appears to favour hydrophobic residues, indicating that Pro277Thr could lead to a major structural rearrangement, likely destabilizing the surrounding beta turn within the DUF domain and perhaps even altering the capacity of the protein to dimerize. Using patient-derived primary fibroblasts, we examined *THAP12* expression at both the RNA and protein levels. Although no significant changes were observed at the transcript level (Fig. 2F, 2G), Western Blot analysis revealed significantly reduced THAP12 protein abundance in both affected patients compared to their unaffected parents (Fig. 2H, 2I). This finding indicates that the *THAP12* variants identified in our patients result in a loss of gene function. In further analysis of the RNA sequencing data from the patient primary fibroblasts, we identified 336 differentially expressed genes (132 downregulated, 204 upregulated, Fig. 2J, 2K and Table S1) between patients and unaffected parents. Gene Ontology analysis revealed dysregulation in several key molecular pathways, including the neuronal system, signal transduction, neurotransmission and GPCR signaling (Fig. 2K). These results suggest that THAP12 could play a role in basic molecular processes and cellular signaling involved in embryogenesis and neurodevelopment.

### Mouse knock-in of patient THAP12 mutations mimics KO embryo lethality

To model *THAP12* variants *in vivo*, we first generated a knockout (KO) allele in mice. Human and mouse *THAP12* share 95% sequence similarity and exhibit highly conserved structural and functional features (Fig. 3A, 3B). Using targeted CRISPR/Cas9 mutagenesis, we established a *Thap12*-KO mouse line carrying a +1bp insertion within the 3^rd^ exon causing a premature stop at position 97 (Fig. 3A, 3C). Intercrosses between heterozygous animals produced no homozygous KO pups among 37 offspring, despite Mendelian inheritance predicting a 25% frequency (Fig. 3E).

**Figure 3.**
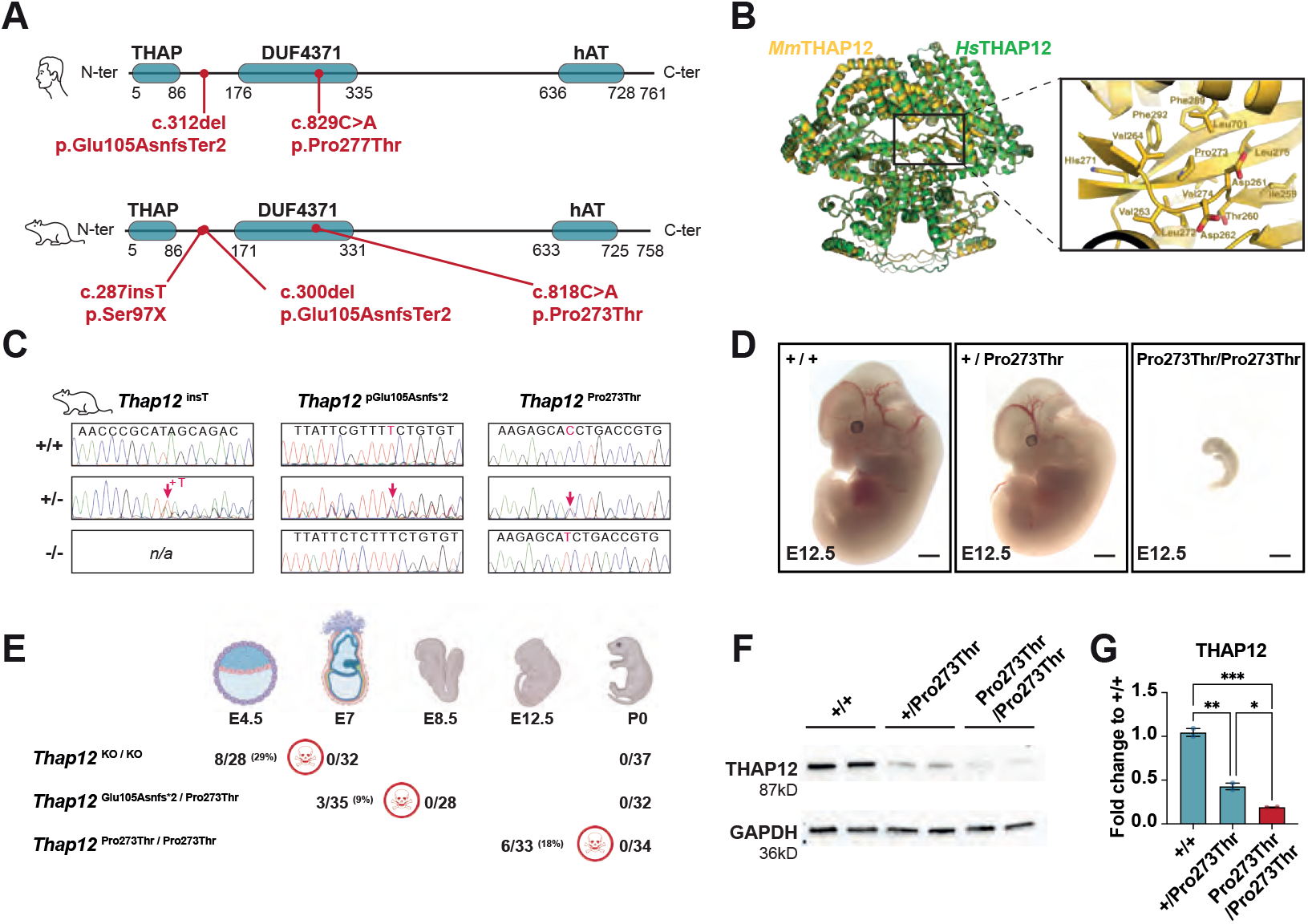
Patient-derived *THAP12* variants are evolutionarily conserved and cause gene loss-of-function and early embryonic lethality in mice. **(A)** Schematic comparison of the human variants identified in the two affected siblings (top) with the corresponding genetically engineered mouse alleles (bottom), showing equivalent positions across conserved domains. **(B)** Structural alignment of the mouse and human THAP12 proteins, whose structures were predicted using AlphaFold3, reveals high overall similarity and highlights the position of the conserved Pro273 residue (orthologous to human Pro277) within a similar hydrophobic core of the DUF4371 domain. **(C)** Sanger sequencing confirms the genotypes of the three engineered alleles in mice (red arrows): *insT, Glu105Asnfs*2* and *Pro273Thr*. **(D)** Representative images of E12.5 mouse embryos show that homozygous *Pro273Thr* mutants are severely developmentally delayed compared to wild-type and heterozygous littermates. **(E)** Table summarizing genotype frequencies at different developmental stages in intercrosses between heterozygous carriers. The absence of homozygous animals at birth confirms early embryonic lethality. Of note, data from both *insT* and *Glu105Asnfs*2* alleles are gathered as “KO” in this panel. Full data for each stage and genotype are provided in Table S2. **(F)** Western blot of E12.5 embryonic heads shows reduced THAP12 protein levels in *Pro273Thr* homozygous mutants. **(G)** Quantification of THAP12 signal relative to wild-type confirms a significant decrease in homozygous embryos. Data are expressed as fold change relative to wild-type levels. *Statistical analyses in panel G used a o*ne-way ANOVA with Tukey post-hoc test: *\*p < 0*.*05, **p < 0*.*01, ***p < 0*.*001*.

The absence of homozygotes indicates that *Thap12* loss-of-function is embryonically lethal, underscoring its essential role during early development *in vivo*. To further investigate the pathogenicity of the *THAP12* variants identified in affected individuals, we generated two knock-in mouse strains carrying each patient-derived mutation by CRISPR-mediated Homology Directed Repair. As noted previously, in human THAP12, the Pro277 residue lies within a hydrophobic core of the DUF domain. The corresponding residue in mouse, Pro273, is similarly embedded in a conserved hydrophobic environment, with complete conservation of the surrounding residues (Fig. 3B). This high degree of structural conservation reinforces the relevance of modelling these exact patients’ variants in mice. We bred both mutant mouse strains in a compound heterozygous configuration, mirroring patients’ genotype. However, no compound heterozygous pups were detected at birth (0/32, Fig. 3E, Table S2). Genotyping of embryos at E7 (early post-implantation gastrula stage) identified a small number of compound heterozygotes (3/35; 9%), but their frequency remained below the expected 25% Mendelian ratio. These results support the pathogenicity of our patient’s *THAP12* variants and indicate that, when modelled in mice, their combined presence is incompatible with early embryo development (Fig. 3E, Table S2).

To dissect the contribution of each variant individually, we also analyzed the effects of the missense mutation (Pro277Thr, at position 273 in mice) in homozygosity. At E12.5, several homozygous *Thap12*^*Pro273Thr/Pro273Thr*^ embryos were recovered at near-Mendelian frequencies (6/33; 18%, Fig. 3E, Table S2). However, these embryos exhibited marked developmental delay compared to wild-type littermates (Fig. 3D). Moreover, Western Blot analysis revealed a significant reduction in THAP12 protein levels in the Pro273Thr homozygous mutants, confirming that the missense variant severely impairs protein stability and results in functional loss of the gene *in vivo* (Fig. 3F, 3G). Together, these *in vivo* data corroborate that our patient-derived *THAP12* mutations lead to gene loss of function. The early embryonic lethality observed in both the KO and knock-in models underscores the essential role of *THAP12* in early murine development.

### Loss of thap12 leads to microcephaly and hypomobility in zebrafish

Given the embryonic lethality observed in our mouse models, we turned to zebrafish as an alternative vertebrate model to gain further insight into THAP12’s developmental functions *in vivo*. Zebrafish possess two *THAP12* orthologs, *thap12a* and *thap12b*, whose proteins share 75% identity with the human THAP12. Structural modelling using Alphafold revealed that both zebrafish orthologs adopt a dimeric configuration highly similar to human *THAP12* (Fig. 4A). As in human *THAP12*, both zebrafish paralogs exhibit predicted monomeric units that comprise an N-terminal zinc-binding domain followed by a domain of unknown functio n (DUF). This high degree of structural conservation supports the relevance of zebrafish as a model for studying *THAP12 function in vivo*.

**Figure 4.**
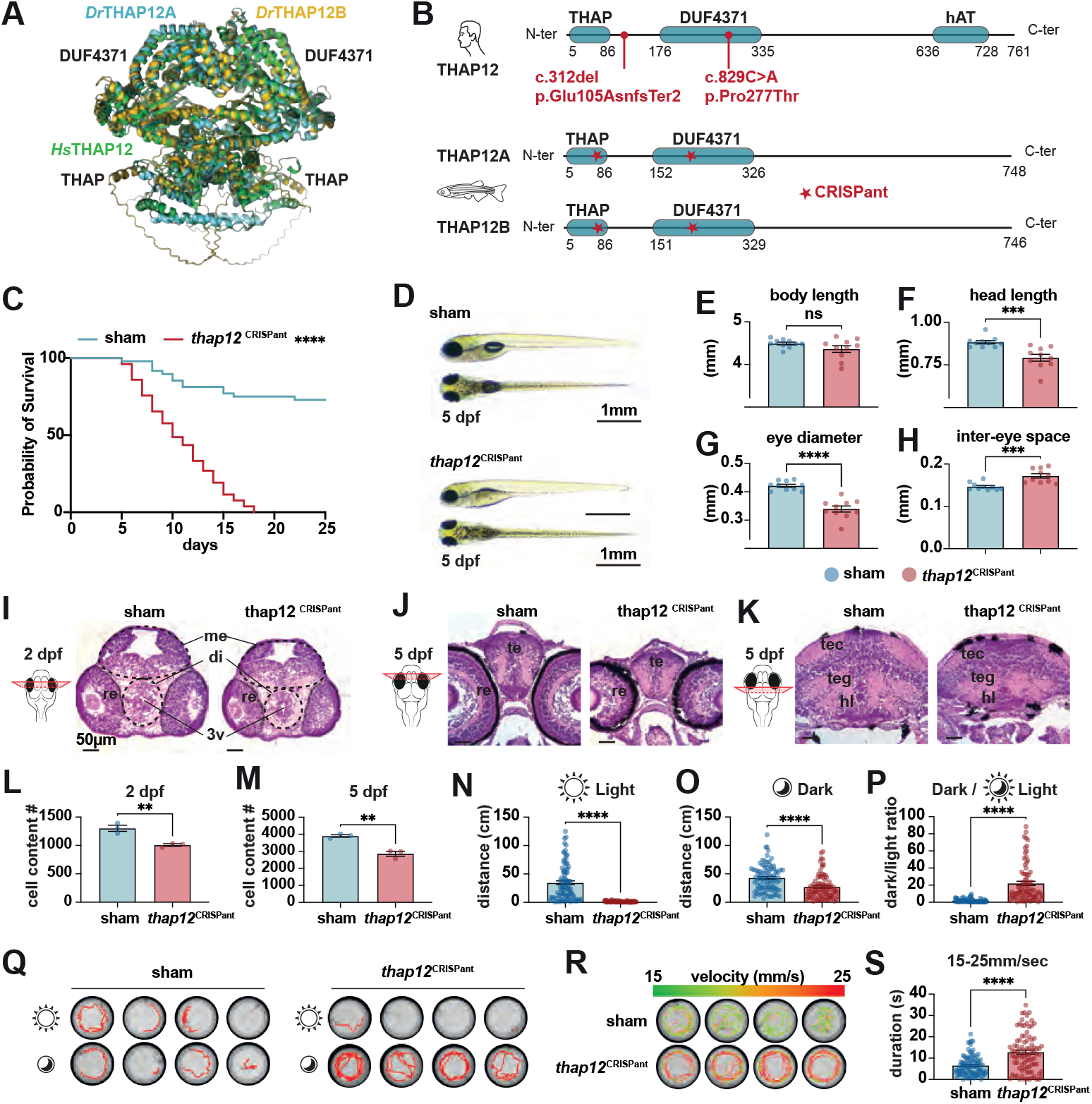
*Thap12 loss* in zebrafish leads to early lethality, microcephaly, and altered behaviour. **(A)** Schematic of the two patient variants and the CRISPR-targeted region in both zebrafish paralogs. **(B)** Structures of human *(Hs)* THAP12, zebrafish *(Dr)*THAP12A, and THAP12B were predicted using AlphaFold3 and their structural alignment reveals strong conservation of the THAP and DUF4371 domains. **(C)** Kaplan-Meier survival curve shows early mortality in *thap12*^CRISPant^ larvae starting around 18 days post-fertilization (**** p<0.0001). **(D)** Lateral views of 5 dpf larvae show reduced head size in *thap12*^CRISPant^ despite preserved body length. **(E-H)** Quantification of body length (**E**), head length (**F**), eye diameter (**G**), and inter-eye distance (**H**) confirms specific morphological defects, including microcephaly, in *thap12*^CRISPant^ larvae. **(I-K)** Haematoxylin and eosin staining of transverse brain sections at 2 dpf (**I**) and 5 dpf (anterior: **J**, posterior: **K**) reveals overall reduced brain size in *thap12*^CRISPant^ larvae (*di: diencephalon, me: mesencephalon, 3v: third ventricule, re: retina, te: telencephalon, tec: tectum; teg: tegmentum; hl: hypothalamus*). **(L-M)** Cell counting from histological sections confirms a significant reduction in brain cell number at both 2 and 5 dpf. **(N-P)** Locomotor assays of 4 dpf larvae show reduced swimming distance under light (**N**) and dark (**O**) conditions, but an overall increased dark/light activity ratio (**P**) in *thap12*^CRISPant^ mutants. **(Q)** Swim tracks (over 30 seconds) illustrating hypoactivity in light and hyperactivity in dark in *thap12*^CRISPant^ at 4 dpf. **(R-S)** Colour-coded swim tracks displaying velocity show a significant increase in high-speed movement (15-25 mm/s) in *thap12*^CRISPant^ compared to sham-injected siblings. *Statistical analyses in panels E-H, L-P and S used unpaired two-tailed Student’s t-test: *p < 0*.*05, **p < 0*.*01, ***p < 0*.*001, ****p < 0*.*0001; ns, not significant*.

To inactivate *thap12* in zebrafish, we used CRISPR/Cas9 to disrupt the expression of both *thap12* orthologs in F0 zebrafish embryos^38^ (Fig. 4B). The mutagenesis efficiency was confirmed by high-resolution melting (HRM) curve analysis and 5-mismatch control gRNAs served as sham-injected controls. (*Fig. S3*). *Thap12* mutant larvae exhibited premature lethality, with no survivors beyond 18 days post-fertilization (dpf) (Fig. 4C). Although overall body development appeared unaffected, mutant larvae failed to inflate their swim bladder by 5 dpf and displayed a reduced head and eye size (Fig. 4D-H). Hematoxylin and eosin staining on transversal brain slices at 2 and 5 dpf revealed a significant decrease in cell density throughout the developing brain of mutant larvae compared to sham-injected siblings (Fig. 4I-M). We assessed motor behaviour in 4 dpf larvae by monitoring spontaneous locomotion under alternating light and dark conditions. Mutant larvae exhibited a general reduction in activity during light phases, but showed increased locomotion in the dark, leading to a significantly increased dark/light motor behaviour ratio (Fig. 4N-Q). This motor phenotype is reminiscent of other zebrafish models of epileptic encephalopathies^39,40^. Particularly, mutants displayed erratic swimming patterns, with rapid, high-amplitude movements during dark phases (Fig. 4R, 4S, video S1), suggesting possible impairments in brain function or neural regulation of behavior.

### Thap12-deficient zebrafish exhibit abnormal neural activity

To further explore the neuronal basis of these behavioral alterations, we used multi-plane two-photon microscopy in Tg(*elavl3*:H2B-GCaMP6s) larvae^41^ to monitor brain-wide nuclear calcium dynamics at single-cell resolution as a proxy for neuronal activity (Fig. 5A, 5B). We recorded 10 minutes of spontaneous brain activity in awake, head-restrained larvae (5-6 dpf, n = 16 larvae per condition; Fig. S4). We aligned each imaging experiment to a larval brain atlas (mapZ ebrain^42^) to compare the activity of multiple brain regions in control and mutant larvae (Fig. 5C). To investigate neuronal communication between brain regions^43^, we computed functional connectivity (FC) matrices from the pairwise correlations of signals averaged within anatomical nodes for each animal (Fig. 5E). Subtracting the group averaged FC matrices of each condition revealed widespread functional changes across the entire brain, with both strengthened and weakened functional connections in mutants (Fig. 5F, 5G). We then used network-based statistics (NBS^44^) to reveal a broadly distributed, significantly altered functional subnetwork, with multiple ventral brain regions exhibiting elevated correlations, and dorsal brain regions such as the habenula and optic tectum exhibiting decreased correlations (Fig. 5H, 5I, P = 0.043, 10,000 permutations). This brain activity phenotype was reproducible, as group-averaged FC differences were significantly correlated across experimental replicates (Fig. S4C-F; Pearson r = 0.353, P = 0.0197, permutation test). Together, these results demonstrate a widespread disorganization of neural activity in *thap12*-mutant zebrafish larvae. Nevertheless, we did not record overall visible generalized seizures or aberrant brain activity patterns, and the overall activity exhibited roughly similar spatiotemporal structure in both CRISPants and sham-injected controls (Fig. 5D). However, when exposed to a low dose of the proconvulsant drug pentylenetetrazole (PTZ), we found that mutant larvae displayed an increased sensitivity, with a significantly increased stereotypical seizure-related motor response^45^ compared to controls, suggesting a lowered threshold for neural hyperexcitability (Fig. 5J, 5K). Taken together, these findings show that *THAP12* loss-of-function in zebrafish leads to early lethality, impaired brain development, and disrupted neural activity patterns. This supports a conserved and essential role for *THAP12* in vertebrate development. Moreover, the behavioural and functional alterations observed in mutant larvae mirror key aspects of the clinical phenotype in our patients, thereby supporting the pathogenicity of *THAP12* loss-of-function.

**Figure 5.**
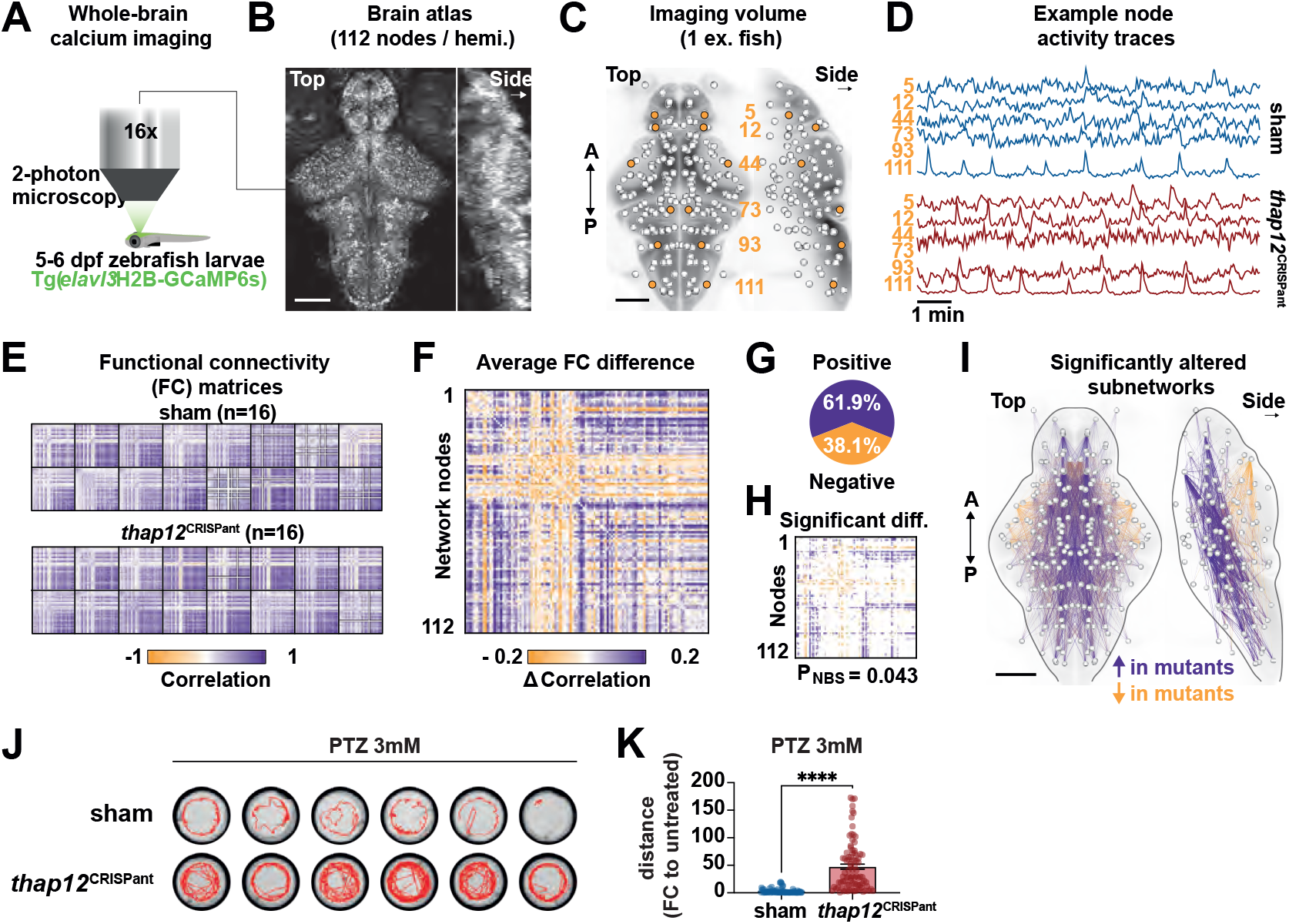
Whole-brain calcium imaging reveals widespread connectivity alterations and seizure susceptibility in *thap12*-deficient zebrafish larvae. (A) Schematic representation of two-photon calcium imaging in *Tg(elavl3:H2B-GCaMP6s)* larvae at 5-6 days post-fertilization. (B) Maximum intensity projection of 21 imaging planes in a control larva. (**C**) 224 network nodes mapped in the mapZebrain larval brain atlas coordinate system; nodes highlighted in orange are associated with the following panel. (**D**) Example node activity traces from 6 anatomical locations in representative larvae from each experimental group; selected nodes correspond to pallium (#5), ventral habenula (#12), optic tectum (#73), cerebellum (#93), medial octavolateralis nucleus (#111), and vagal sensory lobe, respectively. (**E**) Functional Connectivity (FC) matrices from each animal (n=16 per group). (**F**) FC differences obtained by subtracting the group-averaged FC matrices from each other (purple, higher in mutants; yellow, lower in mutants). (**G**) Fractions of positive and negative correlations changes in the previous panel. (**H**) Significantly altered functional network edges (p <0.05); the global significance is assessed using NBS (p = 0.043). (**I**) Anatomical visualization of the significantly altered subnetwork from the previous panel; black outlines roughly indicate the external brain boundaries. (J) Locomotor swim tracks in PTZ-induced seizure-like conditions (3 mM, 10 min) show hyperactivity in *thap12*^CRISPant^. (K) Quantification of distance swum upon 3mM PTZ exposure (fold change to untreated conditions) confirms the hypersensitivity of *thap12*^CRISPant^ to PTZ-induced seizure. *Statistical analyses in panel G-H and K used unpaired two-tailed Student’s t-test: *p < 0*.*05, **p < 0*.*01, ***p < 0*.*001, ****p < 0*.*0001; ns, not significant*.

### Thap12 deficiency alters neurodevelopment through impaired proliferation and increased apoptosis

Our results prompted us to further investigate the nature of the neuronal deficits and the cellular mechanisms affected during brain development. To better characterize the impact of *THAP12* loss of function on neurodevelopment, we generated CRISPant embryos in the Tg[*elavl3*:GFP] transgenic background and measured brain size at 2 and 5 dpf. Mutant larvae displayed a significant reduction in overall brain size at both stages compared to wild-type siblings (Fig. 6A-E and 6G-I), which is consistent with our prior histological observations (Fig. 4F-M). We confirmed that this reduction is associated with a decrease in neuronal density in Elavl3-immunolabeled brain sections (Fig. 6C-E, 6I). Using immunolabeling against acetylated tubulin, which labels axonal tracts, we revealed a disorganized brain structure and a reduced number of axonal projections, particularly within the commissural tracts in *thap12* mutant larvae (Fig. 6F, 6J). Together, these findings point to defects in early neural circuit formation and suggest disrupted neuronal connectivity. To validate the specificity of these CRISPant-induced phenotypes and exclude potential off-target or mosaic effects, we generated stable knockout lines for both *thap12a* and *thap12b orthologs (Fig. S5A-N)*. Homozygous *thap12a*^*-/-*^ mutants recapitulated the major phenotypes observed in CRISPants, including reduced brain size, premature lethality, abnormal light/dark motor behavior, and increased sensitivity to PTZ (*Fig. S5A-N*). In contrast, *thap12b*^*-/-*^ mutants exhibited normal survival and behaviour with no PTZ hypersensitivity, indicating that *thap12a* is the primary ortholog responsible for the zebrafish larval phenotype (Fig. S5C-H). These findings confirm the specificity and robustness of our CRISPant approach. To further dissect the molecular mechanisms underlying the phenotype, and to minimize variability inherent to mosaic editing, we performed bulk RNA sequencing on *thap12a*^*-/-*^ larval brains and identified 287 differentially expressed genes (123 upregulated, 164 downregulated, p.value < 0.05, |logFC| >1, Fig. 6K and Table S3). Gene Ontology (GO) enrichment highlighted key pathways involving p53 signaling path way, cell cycle progression, and metabolic pathways (Fig. 6L). These data suggest that neurodevelopmental defects associated with *thap12* loss may be linked to alterations in cell proliferation and apoptosis. We confirmed this hypothesis experimentally by examining proliferation and apoptosis at 24 hpf, a critical time point for early zebrafish neurogenesis^46,47^. Immunostaining with an anti-phospho-Histone H3 (PH3) antibody, which marks mitotic cells, revealed a significantly reduced number of mitotic cells in the brain region of *thap12* mutants, indicating impaired proliferation (Fig. 6N, 6P). Conversely, Acridine orange and cleaved caspase-3 staining each showed a significantly higher number of apoptotic cells in the developing brains of *thap12* mutants, highlighting increased cell death during early neurogenesis (Fig. 6M, 6O. Fig. S5O, S4P). To confirm the specificity of these phenotypes, we showed that the same changes are observable in *thap12a*^-/-^ mutant compared to their wild-type siblings (Fig. S5Q-S). Moreover, we performed rescue experiments by co-injecting synthetic human *THAP12* wild-type mRNA into *thap12*-mutant embryos. This approach significantly restored the number of PH3-positive mitotic cells at 1 dpf (Fig. 6Q, 6R), confirming that impaired proliferation arises directly from THAP12 deficiency. A partial rescue of brain size was also observed at 2 dpf, although less pronounced, likely due to the transient nature of the injected mRNA (Fig. 6S, 6T). In contrast, co-injection of the patient-derived Pro277Thr variant failed to restore either of those phenotypes (Fig. 6Q-T), providing additional evidence for the pathogenicity of this variant and reinforcing the causal role of THAP12 dysfunction.

**Figure 6.**
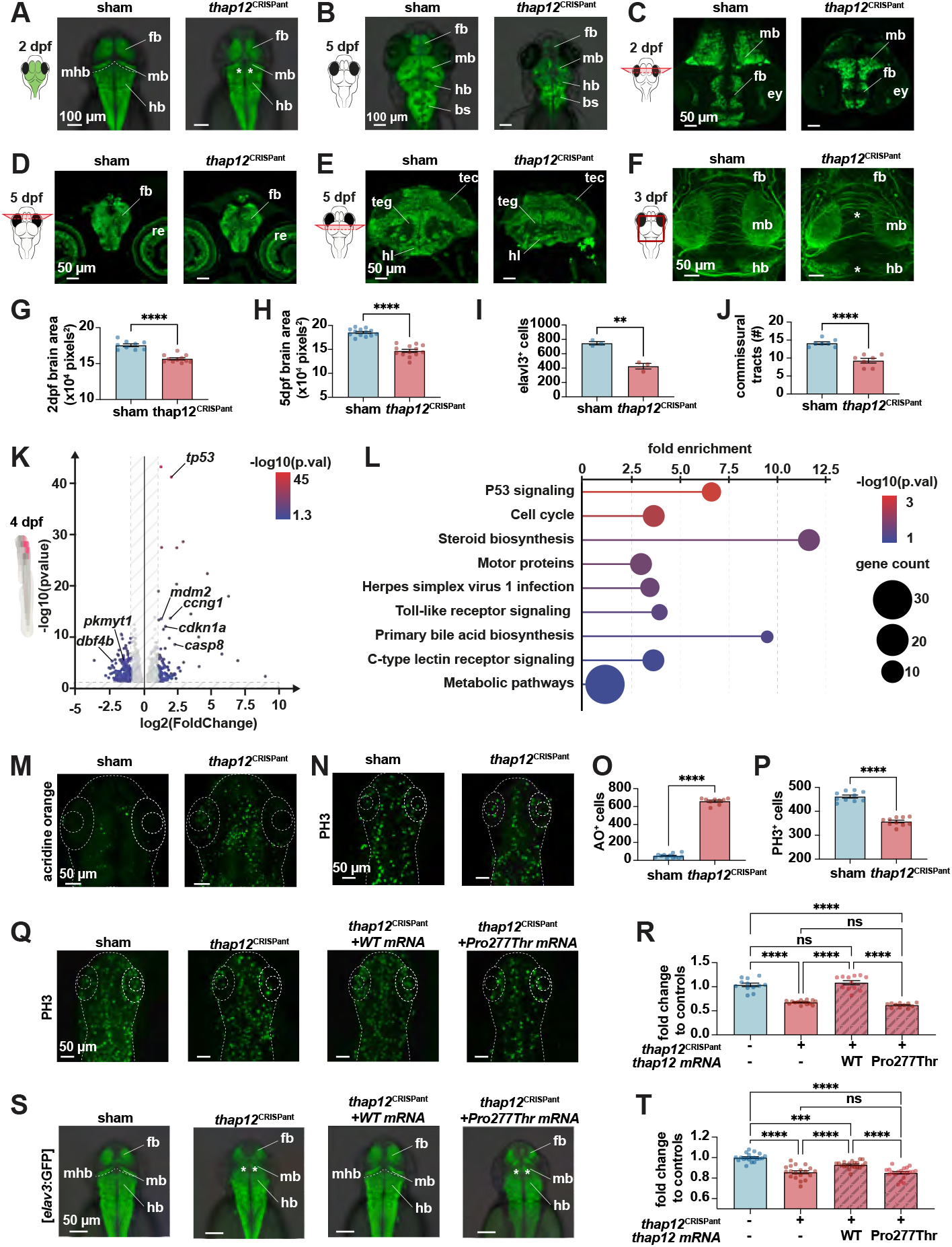
THAP12 loss impairs brain growth and neurogenesis in zebrafish larvae and is rescued by WT mRNA. (A-B) Dorsal views of Tg[*elavl3*:GFP] larvae at 2 dpf (A) and 5 dpf (B) showing a reduced brain size in *thap12*^CRISPant^ compared to sham-injected controls. The lack of a clear midbrain-hindbrain boundary (dotted line) is indicated by asterisks. **(C-E)** Transverse sections immunostained for *elavl3* show reduced brain size in *thap12*^CRISPant^ embryos at 2 dpf (**C**) and at 5 dpf (**D, E**). (F) Dorsal view of 3 dpf brains immunostained for acetylated tubulin showing reduced density of axonal tracts in *thap12*^CRISPant^, especially at the level of the commissure (asterisks). **(G-H)** Quantification of brain area from whole-brain imaging of Tg[elavl3:GFP] larvae at 2 dpf **(G)** and 5 dpf **(H)** confirms a significant reduction of brain size in *thap12*^CRISPant^. **(I-J)** *thap12*^CRISPant^ larvae show reduced numbers of *elavl3*+ neurons from immunolablled cross-sections (I) and commissural axonal tracts **(J). (K)** Volcano plot displaying differentially expressed genes from bulk RNA-sequencing analysis of microdissected larval brains from 4dpf *thap12a*^-/-^ compared to wild-type siblings. Upregulated genes include several involved in apoptosis (e.g. *tp53*) and cell cyle (e.g. *ccng1*). Significance is color-coded according to the -log10(adjusted p-value). **(L)** Pathway enrichment analysis identifies p53 signaling, cell cycle, and metabolic stress as significantly enriched pathways in mutants compared to wild-type siblings. Dot size indicates the number of genes in each pathway; color represents -log10(p-value). **(M)** Acridine orange staining reveals increased cell death in the brain of *thap12*^CRISPant^ at 1 dpf. **(N)** Anti-phospho-H3 immunostaining shows a reduced number of proliferating cells in *thap12*^CRISPRant^ larvae at 1 dpf. Quantification are shown in panel **O** and **P. (Q-T)** Injection of human wild-type *THAP12* mRNA, but not Pro277Thr mutant mRNA, rescues reduced proliferation in *thap12*^CRISPant^ larvae at 1 dpf (Q-R) and partially rescues brain size at 2 dpf **(S-T)**. The lack of a clear midbrain-hindbrain boundary (dotted line) described in panel A is indicated by asterisks. *Scale bars are shown on each panel. Statistical analyses in panel G-J and O-P used unpaired two-tailed Student’s t-test, and in panels R and T used one-way ANOVA: *p < 0*.*05, **p < 0*.*01, ***p < 0*.*001, ****p < 0*.*0001; ns, not significant. fb: forebrain; mb: midbrain; hb: hindbrain; mhb: midbrain-hindbrain boundary (dotted line); bs: brainstem; ey: eye; re: retina; tec: tectum; teg: tegmentum; hl: hypothalamus*.

In conclusion, our results suggest that *THAP12* loss-of-function causes widespread neurodevelopmental defects, including reduced cell proliferation and increased apoptosis during early neurogenesis, resulting in impaired neuronal organization *in vivo*. These cellular and structural abnormalities provide preliminary mechanistic insights into how *THAP12* dysfunction may contribute to the neurodevelopmental phenotype observed in affected individuals.

## Discussion

In this work, we describe two siblings affected by a severe neurodevelopmental disorder, starting with infantile spasms and evolving into Lennox-Gastaut syndrome, both carrying compound heterozygous variants in *THAP12*. Through complementary functional analyses in patient-derived cells, mouse and zebrafish models, we demonstrate that these variants lead to a clear loss of *THAP12* function. Altogether, our data support *THAP12* as a new gene implicated in early-onset epileptic encephalopathies, expanding the list of known genetic causes of these complex disorders. The two variants identified in our patients (e.g., a frameshift and a missense) impact protein function through different mechanisms. The frameshift variant truncates approximately 85% of the C-terminal sequence, including the entire DUF domain (Fig. 2D, 2E), likely abolishing protein dimerization and activity. In line with this, RNA sequencing of patient-derived fibroblasts showed a markedly reduced allelic expression for this variant (Fig. S1C), supporting its classification as a full loss-of-function allele. The Pro277Thr substitution affects a highly conserved hydrophobic motif, likely essential to the structural integrity of the entire DUF domain, potentially leading to protein instability, as observed in our western blot analysis (Fig. 2H, 2I, 3F, 3G). Whether this involves proteasome-related degradation mechanisms remains to be determined. In our study, both parents carry a single functional copy of *THAP12*, supporting the idea that the gene is haplosufficient, i.e., one functional allele is sufficient for normal development. Specifically, the mother carries a frameshift variant predicted to result in a loss-of-function allele, while the father carries a missense variant that we showed leads to markedly reduced protein expression, consistent with a hypomorphic or unstable allele. Interestingly, *THAP12* mRNA levels in the heterozygous mother were not decreased (Fig. 2F), and qPCR analyses in patient-derived fibroblasts even revealed a tendency toward increased transcript levels (Fig. 2G). A similar trend was seen in *thap12* mutant zebrafish, where transcript levels were also significantly elevated (ENSDARG00000042489, log2FC: 0.71, padj: 1.03e-05; Table S3). This suggests the existence of a compensatory mechanism that may upregulate *THAP12* expression when protein levels are reduced, possibly to maintain sufficient levels of functional homodimers.

However, the fact that the function of THAP12 seems to require homodimerization raises the possibility that some heterozygous missense variants could affect normal gene function through dominant-negative mechanisms. In such a scenario, a mutant protein encoded by the altered allele could still dimerize with the wild-type protein but interfere with the dimer’s DNA binding ability or its interaction with cofactors, thus impairing the function of the entire complex. While our study focused on biallelic loss-of-function variants consistent with a recessive inheritance pattern, this hypothesis supports the idea that particular heterozygous missense variants could also be pathogenic despite the presence of a wild-type allele. In contrast, heterozygous null variants such as frameshifts or nonsense mutations may be better tolerated, as the remaining wild-type allele would sufficiently produce fully functional homodimers. This distinction highlights the need for further investigation into the pathogenic potential of rare heterozygous *THAP12* variants in undiagnosed neurodevelopmental disorders

Beyond dominant-negative effects, milder variants (such as hypomorphic or partially destabilizing) may also result in attenuated phenotypes. Because *THAP12* had not been previously linked to neurological diseases, such variants may not have been considered relevant in earlier genetic analyses. Our study encourages re-examining genetic data from less severely affected individuals to assess whether rare *THAP12* variants may be involved. Functional studies of VUS in *THAP12* could be assessed *in vitro* to evaluate their effects on protein expression and stability. Later, as *THAP12* target genes and DNA-binding sites are identified, we will be able to assess how these variants affect its transcriptional activity. Despite extensive outreach through international undiagnosed disease networks, Matchmaker Exchange, and unsolved neurodevelopmental disorder cohorts, no additional individuals carrying biallelic pathogenic variants in *THAP12* were identified. This scarcity likely reflects both the ultra-rare nature of pathogenic variation in *THAP12* and the strong evolutionary constraint acting on the gene. In particular, complete biallelic loss-of-function may be incompatible with survival, requiring a hypomorphic allele with residual function, such as p.Pro277Thr, in trans to a more severe loss-of-function variant to allow viability. This restrictive configuration narrows the mutational spectrum in *THAP12* compatible with disease. The lack of recurrence is not unexpected in the context of ultra-rare disorders and underscores the challenges of gene discovery in the post-genomic era. While the last two decades have seen the identification of hundreds of genes involved in neurological disorders, the remaining ones are likely to be associated with extremely rare variants found in only a few families worldwide. Uncovering their roles requires not only large-scale sequencing efforts but also robust functional validation. Although our study focused on a single family, we believe that the combination of in vitro and in vivo assays mimicking the patient-specific variants, coupled with cDNA rescue experiments, provides compelling evidence supporting the pathogenicity of these ultra-rare *THAP12* variants and their causal role in the disease.

Interestingly, our knock-in mouse models, engineered to carry the exact *THAP12* variants found in patients, showed early embryonic lethality. This phenotype was reminiscent of that of full *Thap12* knockout mice, reinforcing the idea that patient variants result in a complete loss of gene function. The fact that mice die before implantation, whereas human patients survive after birth, probably highlights species-specific differences in how they tolerate THAP12 deficiency, although strain-specific effects, such as those associated with the C57BL/6 genetic background, could also contribute. In contrast to mice, our zebrafish *thap12*-deficient mutants survive up to about two weeks post-fertilization, long enough for brain development and behavioral phenotypes to be fully assessed. These larvae show clear signs of microcephaly, abnormal locomotion, increased seizure susceptibility and abnormal brain activity. Recent comparative transcriptomic data suggest that the developmental staging of a 6-day-old zebrafish larva roughly corresponds to that of a late-prenatal human brain^48^, which is consistent with the clinical presentation observed in our patients. For future preclinical studies investigating *THAP12*-related brain diseases, it will be worth noting that the zebrafish may offer unique advantages over mouse models for recapitulating early neurodevelopmental defects, as it is the case for other epileptic encephalopathy syndromes such as Dravet syndrome^49^. Importantly, the scope of our findings goes beyond the diagnosis of a rare disease. This is the first study to associate *THAP12* with a neurological phenotype. *THAP12* is part of the Thanatos-Associated Protein (THAP) family, whose members share a conserved DNA-binding domain and are known to regulate gene transcription^18-20^, cell proliferation^21-24^, mitochondrial function^23,29,50^, and apoptosis^25-28^. Some, such as *THAP11*, have already been implicated in neurogenesis in model systems and linked to brain disorders in humans^51-53^. Our work now positions *THAP12* in this landscape and suggests that other THAP family members may also play roles in brain disorders. While we focused here on the consequences of *THAP12* loss-of-function during brain development, its exact molecular role remains to be defined. Previous studies under alternative aliases such as *PRKRIR* or *DAP4* have suggested roles in apoptosis and various signal transduction pathways^25,54^. However, these findings were based on different *in vitro* models and do not clarify its function in the developing brain. Notably, although *THAP12* shares the highly conserved THAP-type zinc finger domain found across the gene family, the ability of endogenous THAP12 to bind DNA and regulate gene expression has not been experimentally tested. Understanding these molecular mechanisms is now essential. It will not only shed light on how *THAP12* loss leads to disease but also help identify potential downstream targets relevant to future therapeutic strategies.

## Methods

### Primary fibroblasts culture

Primary fibroblasts from patients were ordered from Coriell (IDs GM27990, GM27993, GM27995, and GM27997) and cultured according to the supplier’s guidelines. Briefly, cells were cultured in Dulbecco’s Modified Eagle Medium/Nutrient Mixture F-12 (DMEM/F12, Gibco) supplemented with 15% fetal bovine serum (FBS) and 1% penicillin/streptomycin (100 U/mL). Fibroblasts were maintained in a humidified incubator at 37°C with 5% CO2. Cells were passaged at 80-90% confluence using 0.25% trypsin-EDTA, and passages 5-8 were used for all experiments.

### Genomic DNA extraction

Genomic DNA was extracted from approximately 5×10^6^ primary fibroblasts after the 5^th^ passage using the MyTaq Extract-PCR Kit (BIO-21126, meridian bioscience) following the manufacturers’ protocol.

### Whole Genome Sequencing and RNA sequencing form primary fibroblasts

Genome sequencing and data processing were performed by the Genomics Platform at the Broad Institute of MIT and Harvard. PCR-free preparation of sample DNA (350 ng input at >2 ng/ul) is accomplished using Illumina HiSeq X Ten v2 chemistry. Libraries are sequenced to a mean target coverage of >30x. Total RNA was isolated from fibroblasts using the RNeasy Plus Mini Kit (Qiagen) according to the manufacturer’s protocol. Total RNA was quantified using the Quant-iT™ RiboGreen® RNA Assay Kit and normalized to 5ng/ul. Flowcell cluster amplification and sequencing were performed according to the manufacturer’s protocols using the Illumina NovaSeq. Each run was a 101bp paired-end with an eight-base index barcode read.

Whole-genome sequencing (WGS) and RNA-seq data were analyzed using separate pipelines. For WGS, FASTQ files were aligned to the human hg19 reference genome using BWA, and variants were called using GATK HaplotypeCaller. RNA-seq reads were aligned to hg19 using HISAT2, and gene expression quantification was performed with HTSeq-Count. Annotation was performed using ANNOVAR and custom scripts, generating a VCF file for each individual. Variant prioritization focused on rare variants shared by both affected individuals and inherited from either parent. Filtering included frequency thresholds and pathogenicity metrics (GERP > 4, PhastCons > 400, SIFT > 0.5, PolyPhen-2 > 0.95, CADD > 15). Variants were visualized in IGV, and candidate splicing alterations were assessed using SpliceAI. Differential expression analysis was conducted on RNA-seq data using DESeq2. Shared variants from both probands were extracted, and a recessive inheritance model was applied to prioritize homozygous or compound heterozygous variants.

### Quantitative PCR

Total RNA was extracted from either patient-derived fibroblasts or whole zebrafish larvae using the RNeasy Plus Mini Kit (Qiagen), following the manufacturer’s protocol. cDNA synthesis was performed using 500 ng of total RNA with the SuperScript VILO cDNA Synthesis Kit (Invitrogen). The resulting cDNA was diluted 1:20 and subjected to quantitative PCR using SYBR Green I Master mix (Roche) on a LightCycler 96 system (Roche). Gene expression was normalized to *GAPDH* and *ACTB*, and relative quantification was calculated using the 2^-ΔΔCt^ method^55^. Primer sequences are available in Table S4.

### Primary fibroblast protein extraction and Western blotting

Primary fibroblasts were harvested at 80–90% confluency, washed with cold PBS, and detached using 0.25% trypsin at 37 °C for 3 minutes. Cells were pelleted by centrifugation at 1000 rcf for 5 minutes and flash frozen. Cell pellets were lysed in RIPA buffer (Sigma Millipore) supplemented with protease inhibitor cocktail (Sigma Millipore) and phosphatase inhibitor cocktail (PhosSTOP, Roche Diagnostics). Lysates were sonicated on ice twice for 30 seconds at 50% amplitude, with a 10-second pause between pulses. Clarified lysates were quantified using the DC Protein Assay (Bio-Rad). 30µg of total protein were mixed with Laemmli buffer, denatured at 95 °C for 10 minutes, resolved by SDS-PAGE on 4–20% Mini-PROTEAN TGX Precast Gels (Bio-Rad), and transferred to nitrocellulose membranes. Membranes were blocked in TBS-T with 5% milk for 1 hour at room temperature and incubated overnight at 4 °C with the following primary antibodies: anti-THAP12 (1:500, Invitrogen, PA5-112672) and anti-GAPDH (1:2000, Cell Signaling, 2118S). After washing, membranes were incubated with HRP-conjugated goat anti-rabbit IgG secondary antibody (1:2000–1:5000) for 1 hour at room temperature. Detection was performed using enhanced chemiluminescence, and bands were visualized with a ChemiDoc MP imaging system (Bio-Rad). Densitometry was performed using Image Laboratory software v5.0 (Bio-Rad).

### Structure Prediction Modelling

Structures of human THAP12 (Uniprot accession O43422), mouse THAP12 (Uniprot accession Q9CUX1), zebrafish THAP12a (Uniprot accession Q803J5) and THAP12b (Uniprot accession Q6DI10) were predicted with AlphaFold3 (alphafoldserver.com) using two copies of each sequence as prediction inputs. As the exact DNA sequences bound by THAP12 still remain elusive, modeling human THAP12 in complex with DNA was performed using two copies of human THAP12 along with random DNA sequences GAACATGTCCCAACATGTTT and AAACATGTTGGGACATGTTC (reverse complement). PyMOL (pymol.org) was used for structure visualization, structural alignments, and figure preparation.

### Mice Housing

C57BL/6J mice (Jackson Laboratory) were used as embryo donors and CD-1 females (Charles River) served as pseudopregnant recipients. All procedures were approved by the CRCHUM Animal Care Committee in accordance with Canadian Council on Animal Care (CCAC) guidelines. Animals were housed under standard SPF conditions with a 12-hour light/dark cycle and ad libitum access to food and water.

### Mouse Genetic Engineering

CRISPR reagents included recombinant Cas9 protein (IDT), synthetic Alt-R™ crRNA and tracrRNA (IDT). Guide RNAs (gRNAs) were annealed by incubating equimolar crRNA and tracrRNA at 95°C for 5 minutes and cooling to room temperature. Cas9 ribonucleoprotein (RNP) complexes were assembled by incubating 80 µM Cas9 with 80 µM pgRNA for 10 minutes, followed by dilution in 20 µL of Opti-MEM (ThermoFisher, #31985070) containing 10 µM of single-stranded donor DNA (ssODN; Ultramer, IDT). Due to embryonic lethality observed in direct knock-in attempts, we implemented a two-step strategy. In a first round, a silent mutation disrupting the protospacer adjacent motif (PAM) site was introduced via CRISPR knock-in using a dedicated gRNA/ssODN pair. F0 mosaic animals were screened by Sanger sequencing (ear punches at P21), and germline-transmitting heterozygotes were crossed to WT to establish stable PAM-mutated heterozygous lines.

These were used for a second CRISPR-editing round targeting the remaining wildtype PAM allele with ssODN encoding the patient-specific variant along surrounding silent mutations. In total, two distinct gRNA/ssODN combinations per line were used, generating four lines in parallel. gRNA and ssODN sequences are provided in Table S4. One-cell embryos were collected from superovulated 3-week-old C57BL/6J females (5 IU PMSG + 5 IU hCG) and kept in EmbryoMax KSOM medium (Sigma). Electroporation was performed in 1 mm cuvettes (BioRad) using a Gene Pulser XCell (30 V, 3 ms, 2 pulses, 100 ms interval). Surviving embryos were transferred into pseudopregnant CD-1 females at 0.5 dpc. Ear biopsies from 21-day-old pups were genotyped using the MyTaq Extract PCR kit (Bioline). PCR products were amplified with Q5 High-Fidelity polymerase (NEB) using locus-specific primers (see Table S4) and sequenced at the Genome Québec platform. Alleles were confirmed by alignment in SnapGene.

### Mouse embryo dissection and Western Blot

E12.5 embryos were dissected in phosphate-buffered saline (PBS). Heads were collected from wild-type and heterozygous embryos, while whole embryos were used for homozygous samples due to severe developmental delay. Genotyping was performed prior to protein extraction. Tissues were lysed in RIPA buffer (Sigma Millipore) supplemented with protease (Sigma Millipore) and phosphatase inhibitors (PhosSTOP, Roche Diagnostics), and sonicated on ice (2×30 s, 50% amplitude, 10 s rest). Protein lysates were quantified using the DC Protein Assay Kit (Bio-Rad). Lysates were mixed with Laemmli buffer, denatured (95°C, 10min), and 50 µg per lane were separated on 4–20% Mini-PROTEAN TGX Precast Gels (Bio-Rad), then transferred to nitrocellulose membranes. Membranes were blocked (TBS, 0.1% Tween 20, 5% skim milk, 1 h, RT) and incubated overnight at 4°C with anti-THAP12 (1:2000, Bethyl Laboratories, A300-586A) and anti-GAPDH (1:2000, Invitrogen, MA5-44678). After washes, membranes were incubated with HRP-conjugated goat anti-rabbit IgG (1:10,000, Sigma Millipore, 12-348) for 1 h at RT. Detection was performed using ECL, and imaging and densitometry were conducted with the ChemiDoc MP and Image Lab software v5.0 (Bio-Rad).

### Zebrafish Housing

Zebrafish (*Danio rerio*) maintenance was performed following standard procedures (Westerfield, 2007) at 28.5°C under a 12-hour light/12-hour dark photoperiod at the animal facility of the University of Montreal Hospital Research Center (CRCHUM), Montreal, QC, Canada. All experiments were conducted in accordance with the guidelines of the Canadian Council on Animal Care and approved by the institutional animal care committee.

### Zebrafish F0-CRISPant model generation

Zebrafish-optimized Cas9 mRNA was synthesized in vitro using the mMESSAGE mMACHINE SP6 Transcription Kit (Ambion) and a NotI-linearized pCS2-nCas9n plasmid (Addgene #47929). Four guide RNAs (gRNAs) targeting *thap12a* and *thap12b* were designed with CRISPRscan and synthesized by Synthego. gRNA sequences are provided in Table S4. The gRNA sequences were: *thap12a* exon 2: 5′ GGGUACUUGUUCAGAUGAUC 3′ ; *thap12a* exon 5: 5′ GGGGGUUGACAGCAGCCCUC 3′; *thap12b* exon 3: 5′ GGGGGGGUUGCCUAGAUGAC 3′; *thap12b* exon 5: 5′ GGUGAGCCGAGGUACAUCAU 3′. At the one-cell stage, wild-type *Tübingen long fin* (TL) embryos were injected with 1 nL of a CRISPR mix containing 100ng/µL Cas9 mRNA and 30 ng/µL pooled gRNAs, using a Picospritzer III pressure microinjector (Parker Hannifin).

### Zebrafish stable CRISPR line generation

To generate stable mutant lines, we selected the most efficient gRNAs targeting exon 2 of *thap12a* and exon 3 of *thap12b*. Founder F0 fish exhibiting high mutation efficiency at the respective loci, as confirmed by HRM analysis and Sanger sequencing, were outcrossed to wild-type Tübingen long fin (TL) fish. F1 progeny were screened for germline transmission of indels causing frameshift mutations and premature stop codons. Confirmed heterozygous carriers were incrossed to obtain homozygous F2 embryos, which were used for phenotypic and molecular characterization.

### Zebrafish genotyping

Genomic DNA was extracted from individual embryos or tail fin clips using alkaline lysis as previously described^56^. High-Resolution Melting (HRM) analysis was performed using a LightCycler 96 instrument (Roche) under standard HRM cycling conditions. Genotypes were inferred from altered melting profiles relative to wild-type controls using LightCycler 96 software v1.1 (Roche). The same gDNA was also used for conventional PCR amplification using standard protocols and a Bio-Rad thermocycler. PCR products spanning the target sites were submitted for Sanger sequencing (Genome QC) to precisely characterize the indel mutations and confirm frameshift alleles. All primer sequences flanking each CRISPR target site are listed in Table S4.

### Zebrafish larval survival assay

Approximately 30 embryos per condition were raised in glass beakers containing 300 mL of E3 medium and maintained at 28 °C under a 12 h light/12 h dark photoperiod. Survival was monitored across at least two independent experiments, with a minimum of 30 larvae per batch. Survival data were analyzed using Kaplan–Meier survival curves in GraphPad Prism version 10.

### Zebrafish larvae morphology measurements

At 5 dpf, larvae were anesthetized in 4% buffered tricaine (MS-222) and embedded in 2% methylcellulose for imaging. Dorsal and lateral images were acquired at 4× magnification using a Leica stereomicroscope. Morphometric measurements were performed manually in ImageJ (version 2.3.0) and included body length (anterior tip of the head to the posterior tip of the tail), eye diameter (widest diagonal across the eye in lateral view), and head dimensions (length from snout to hindbrain and width at the widest point in dorsal view).

### Hematoxylin and Eosin (H&E) Staining

Whole larvae at 2 and 5 dpf were fixed, dehydrated, and embedded in paraffin. Transverse sections (5 µm) were obtained using a Leica Jung Biocut microtome. Sections were deparaffinized, rehydrated, and stained with hematoxylin for 4 min, differentiated in acid–alcohol, rinsed in tap water, and incubated in lithium carbonate for 10 s. Slides were counterstained with eosin for 2 min, dehydrated through graded alcohols, cleared in xylene, and mounted using Permount (Sigma Millipore). Images were captured at 40× magnification using a Leica brightfield microscope. Quantification was performed in ImageJ (version 2.3.0). At 2 dpf, cells were counted in the head (excluding eyes), and at 5 dpf, counts were performed in the optic tectum.

### Zebrafish larvae swimming assay

At 4 dpf, individual larvae were transferred to 96-well plates containing 200 µL of E3 medium per well. Plates were placed in a DanioVision™ observation chamber (Noldus Information Technology), and larvae were acclimated in the dark for 30 min prior to recording. Locomotor activity was recorded during alternating 1-hour light and dark phases. Total swimming distance was quantified using EthoVision XT 13 software (Noldus). Representative swim trajectories were extracted using the integrated track visualization tool in EthoVision.

### Confocal live transgenic imaging

*Tg(elavl3:GFP)* embryos were raised in E3 medium with 0.003% 1-phenyl-2-thiourea (PTU; Sigma Millipore) from 24 hpf to prevent pigmentation. At 2 and 5 dpf, larvae were anesthetized with 4% buffered tricaine (MS-222) and mounted in 1.5% low-melting point agarose (Thermo Scientific). Z-stack images were acquired at 10× magnification with 2 µm steps using an Olympus confocal microscope. Image processing was performed in Volocity (Quorum Technologies), and GFP-positive brain area was quantified using ImageJ (v2.3.0).

### Zebrafish larvae in toto immunofluorescence

For mitotic labeling, 1 dpf larvae were fixed in 4% paraformaldehyde (PFA, Thermo Scientific) for 1 hour at 4°C, washed in 1% Triton X-100 in PBS, permeabilized in 100% acetone (10 min, -20°C), and blocked in 5% normal goat serum (NGS, Jackson ImmunoResearch), 2% BSA, and 1% Triton in PBS. Larvae were incubated overnight at 4°C with anti-phospho-histone H3 (Millipore #09-838; 1:750). Alexa Fluor 488-conjugated goat anti-rabbit secondary antibody (Millipore # A11034; 1:1000) was used for detection. For apoptosis detection, larvae were fixed overnight in 4% PFA, permeabilized in 1% Triton X-100, blocked in 10% NGS, 2% BSA, and incubated overnight at 4°C with anti-cleaved caspase-3 (Cell Signaling #9661; 1:200). Detection was performed as above. Confocal imaging was done at 10× magnification (Olympus confocal microscope). Positive cells were counted in ImageJ. For acetylated tubulin labelling, larvae were fixed overnight at 4°C in Dent’s fixative (80% methanol, 20% DMSO), rehydrated through a graded methanol series, and washed in PBS-Tween (0.1% Tween-20 in PBS). After 1 hour blocking in 1% Tween-20, 2% BSA, and 10% NGS in PBS, larvae were incubated overnight at 4°C with anti-acetylated α-tubulin (Sigma Millipore #T7451; 1:500). Alexa Fluor 488-conjugated goat anti-mouse secondary antibody (Sigma Millipore # A11029; 1:1000) was applied the following day. Larvae were mounted in agarose for imaging and processed as above.

### Acridine orange live staining

1 dpf larvae were incubated in 10 µg/mL acridine orange (Sigma Millipore) in E3 medium for 20–30 min at room temperature. Larvae were washed in PBS, anesthetized with 4% tricaine, and mounted in agarose. Fluorescent signals were imaged using Olympus confocal microscopy and manually quantified in ImageJ.

### Zebrafish larval brain cryosections and immunolabelling

Larvae were fixed in 4% PFA, cryoprotected in 15% then 30% sucrose in PBS at 4°C, embedded in OCT (VWR), and frozen at -80°C. Transverse cryosections (10 µm at 2 dpf, 15 µm at 5 dpf) were cut throughout the brain. Slides were washed in PBS-Triton (0.1%), blocked with 5% NGS in PBS-Triton, and incubated overnight with anti-HuC/HuD (Elavl3/4; Thermo Fisher # A-21271; 1:500). Alexa Fluor 488 goat anti-mouse secondary antibody (Sigma Millipore #A11029; 1:1000) was applied, and sections were mounted with Fluoromount-G (Invitrogen). Imaging was performed at 20× using Zeiss Apotome.2 and ZEN software.

### Rescue experiments with human THAP12 mRNA

Full-length human THAP12 cDNA was amplified from human fibroblast cDNA and cloned into the pCS2+ vector. Site-directed mutagenesis was performed on the wild-type THAP12 cDNA using primers encompassing the c.829C>A (Pro277Thr) variant to generate the mutant construct (Table S1). pCS2+-THAP12 vectors were linearized, and capped mRNA was synthesized using the mMESSAGE mMACHINE kit (Ambion), with quality verified by agarose gel electrophoresis. One-cell stage wild-type zebrafish embryos were co-injected with 100 ng/µL Cas9 mRNA, 30 ng/µL gRNA mix, and 300 ng/µL human THAP12 mRNA (wild-type or Pro277Thr variant). Control groups included CRISPR-only and mismatch-injected embryos. Phenotypic rescue was assessed by phospho-histone H3 immunostaining and in toto imaging using the Tg(*elavl3*:GFP) line as previously described.

### Zebrafish brain RNA sequencing

Larval brains were microdissected from 4 dpf larvae using fine forceps in calcium-free Ringers’ solution. RNA was extracted from pools of four to five pre-genotyped larvae per condition using the picopure RNA extraction kit (ThermoFisher). Libraries were prepared using the NEBNext dual index kit and sequenced (100 bp paired-end) on an Illumina NovaSeq 6000 platform, producing ∼25M reads/sample. Reads were quality-checked and trimmed with Trimmomatic (v0.39), aligned to the *Danio rerio* GRCz11 genome using STAR (v2.7.9a) in two-pass mode, and quantified with Salmon (v1.4.0). Differential expression was analyzed in R (v4.3.1) with DESeq2 (v1.42.1), using Ensembl v104 annotations improved with RefSeq.

### Zebrafish larvae calcium imaging

CRISPR and sham-injected zebrafish larvae (4 dpf, *Tg(elavl3:H2B-GCaMP6s)*) were embedded in 2% low-melting point agarose in glass-bottom petri dishes. Agarose was removed around the tail using a scalpel, and the preparation was submerged in E3 medium. Spontaneous brain-wide calcium activity was recorded in the dark for 10 minutes using resonant-scanning two-photon microscopy at an excitation wavelength of 920 nm. Images were acquired with a 16× water-dipping objective (NA 0.8), and a piezo-driven objective mount enabled sequential acquisition of 22 imaging planes spaced approximately 12 µm apart, at a volumetric rate of 1 Hz. Prior to functional imaging, a high-resolution anatomical stack was acquired at the isosbestic excitation wavelength of 860 nm (2 µm plane spacing, 24 averaged frames per plane), and used for anatomical alignment of the calcium imaging volume. Imaging planes were corrected for lateral (XY) drift using the NoRMCorre algorithm. Fluorescent nuclei were segmented using a custom local maxima detection algorithm, with each plane treated independently due to the non-overlapping acquisition. Fluorescence traces were extracted for each detected neuron, and activity from all 22 planes was reconstituted into a 3D volume, typically representing ∼50,000 neurons per animal distributed across most brain regions. To map neurons to anatomical space, the 920 nm functional volume was non-rigidly registered to the corresponding 860 nm anatomical volume using the SyN algorithm (Advanced Normalization Tools). The 860 nm stack was then registered to a *Tg(elavl3:H2B-GCaMP6s)* template brain from the mapZebrain 4 dpf larval zebrafish atlas. Neuron XYZ coordinates were transformed into atlas space and assigned to one of 65 non-overlapping anatomical brain regions. For each larva, the activity of neurons within each region was averaged, and regional traces were correlated to generate a functional connectivity (FC) matrix describing pairwise interactions. To identify alterations in functional brain networks Network-Based Statistics (NBS) was used. Standard *t*-tests were applied to each network edge, and edges with *P* < 0.05 were retained. To correct for multiple comparisons, the size of the largest connected component among significant edges was computed and compared to a null distribution generated by group-label shuffling and repeated testing.

### PTZ-induced zebrafish hyperactivity assay

At 4 days post-fertilization (dpf), individual larvae were transferred into separate wells of a flat-bottom 96-well plate (200 µL total volume per well), each initially filled with 180 µL of E3 medium. The plate was placed inside a DanioVision™ observation chamber (Noldus Information Technology), and baseline locomotor activity was recorded for 1 hour under constant light conditions. After this period, 20 µL of a 30 mM pentylenetetrazole (PTZ) stock solution (Sigma Millipore) was added to each well to reach a final concentration of 3 mM. Larvae were then recorded for an additional hour under the same lighting conditions to assess PTZ-induced behavioral responses. Swimming trajectories and locomotor activity were quantified using EthoVision XT 13 software (Noldus Information Technology).

## Supporting information

supplemental informations

## Data Availability

All data produced in the present study are available upon reasonable request to the authors

## Acknowledgment

We thank the CRCHUM animal facility and the genetic engineering and animal modelling platform for their help and support in generating and maintaining our mice and fish colonies. A.B. was supported in part by PROTEO, a Fonds de recherche du Québec - Nature et technologie (FRQNT)-funded network. This work is also supported by a grant from the CERMO-FC to E.S., M.T. and L.C. A.L. was supported by a studentship from the Natural Sciences and Engineering Research Council of Canada (NSERC). S.A. was supported by a Canadian Institute of Health Research doctoral fellowship. M.T. and E.S received a salary award from the Fond de Recherche du Quebec Santé. Funding for P.D.K. and P.D. was from NSERC discovery grants. Funding for M.T. was from Courtois Foundation. Funding for E.S. was from the Mirella and Lino Saputo Foundation, the Lightning and Love Foundation, Brain Canada and the Rare disease models and mechanisms network. Whole genome sequencing and analysis and RNA sequencing data were provided the Broad Institute Center for Mendelian Genomics (CMG) and were funded by the National Human Genome Research Institute (NHGRI) grants UM1HG008900 (with additional support from the National Eye Institute, and the National Heart, Lung and Blood Institute), R01HG009141, U01HG011755, and in part by the Chan Zuckerberg Initiative Donor-Advised Fund at the Silicon Valley Community Foundation grants 2019-199278, 2020-224274 (https://doi.org/10.37921/236582yuakxy) (funder DOI 10.13039/100014989). The content is solely the responsibility of the authors and does not necessarily represent the official views of the funding agencies.

## Authors Contributions

Conceptualization: E.S., M.G. Methodology: K.O., B.R., A.L., A.D.S.B., M.G. Investigation: K.O., B.R., V.T., S.A., A.B., G.B., M.L., J.P., M.G., L.P., G.V., E.S. Formal Analysis and Data Curation: K.O., B.R., V.T., S.A., A.B., G.B., M.L., J.P., M.G., L.P. Writing – Original Draft: K.O., E.S. Writing – Review and Editing: A.L., A.O.-L., D.W., L.C. Visualization: A.L., A.B., L.C., E.S. Supervision: A.O.-L., N.L., J.-F.S., L.C., P.D., P.D.D., M.T., E.S. Project Administration: E.S. Funding Acquisition: L.C., P.D., P.D.D., M.T., E.S.

## Human Data Ethical Approval and Data Access

The IRB of Massachusetts General Brigham (Protocol# 2016P001422) gave ethical approval for the collection of human samples and data analysis. Genome sequencing, RNA sequencing, and phenotypic data from the Broad CMG are available as part of the GREGoR consortium cohort via dbGaP accession numbers phs003047.v4.p3 as RGP_431. Access is managed by a data access committee designated by dbGaP and is based on the intended use of the requester and allowed use of the data submitter as defined by consent codes.

