## supplemental informations for "Ultra-rare biallelic *THAP12* variants cause loss of function and underlie severe epileptic encephalopathy"

**Supplementary Figure S1. Whole-genome sequencing identifies compound heterozygous *THAP12* variants in two affected siblings. (A)** Pedigree and segregation of the candidate *THAP12* variants. Both affected individuals carry a maternally inherited frameshift variant (c.312del, p.Glu105Asnfs*2) and a paternally inherited missense variant (c.829C>A, p.Pro277Thr). **(B-C)** Read-level confirmation of each *THAP12* variant in RNA-seq and WGS data from patients' primary fibroblats.

**A**

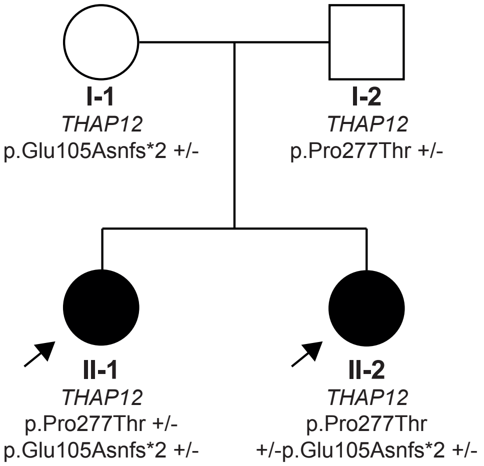


**B** **Missense variant (chr11:76063365 (G>T)) analysis (WGS and RNA-seq)**

| Individual | RNA-seq Coverage | RNA-seq Allelic Ratio (T/(G + T)) | WGS Coverage (G:T) | WGS Allelic Ratio Ratio (T/(G + T)) | Variant |
|---|---|---|---|---|---|
| Mother | G:107 | 0 / (107 + 0) = 0.00 | G:34 | 0 / (34 + 0) = 0.00 | Hom (G:G) |
| Father | G:40; T:60 | 60 / (40 + 60) = 0.60 | G:14; T:20 | 20 / (14 + 20) = 0.59 | Het (G:T) |
| Proband 1 | G:50; T:26 | 26 / (50 + 26) = 0.34 | G:15; T:25 | 25 / (15 + 25) = 0.62 | Het (G:T) |
| Proband 2 | G:39; T:28 | 28 / (39 + 28) = 0.42 | G:23; T:21 | 21 / (23 + 21) = 0.48 | Het (G:T) |

**C** **Frameshift variant (chr11:76072005 (CT>C)) analysis (WGS and RNA-seq)**

| Individual | RNA-seq Coverage | RNA-seq Allelic Ratio (DEL:T) | WGS Coverage | WGS Allelic Ratio Ratio (DEL:T) | Variant |
|---|---|---|---|---|---|
| Mother | DEL:11; T:66 | 11 / (11 + 66) = 0.14 | DEL:16; T:16 | 16 / (16 + 16) = 0.50 | Het(DEL:T) |
| Father | DEL:0; T:100 | 0 / (0 + 100) = 0.00 | DEL:0; T:21 | 0 / (0 + 21) = 0.00 | Hom (T:T) |
| Proband 1 | DEL:7; T:32 | 7 / (7 + 32) = 0.18 | DEL:19; T:11 | 19 / (19 + 11) = 0.63 | Het (DEL:T) |
| Proband 2 | DEL:6; T:35 | 6 / (6 + 35) = 0.15 | DEL:19; T:19 | 19 / (19 + 19) = 0.50 | Het (DEL:T) |

**Supplementary Figure S2. Structural comparison of the predicted structure of THAP12 in absence and presence of DNA.** Human THAP12 was modeled as a dimer in **(A)** absence or **(B)** presence of DNA. Whereas the DUF domains remain largely unaffected, DNA binding triggers a change in orientation of the THAP domains.

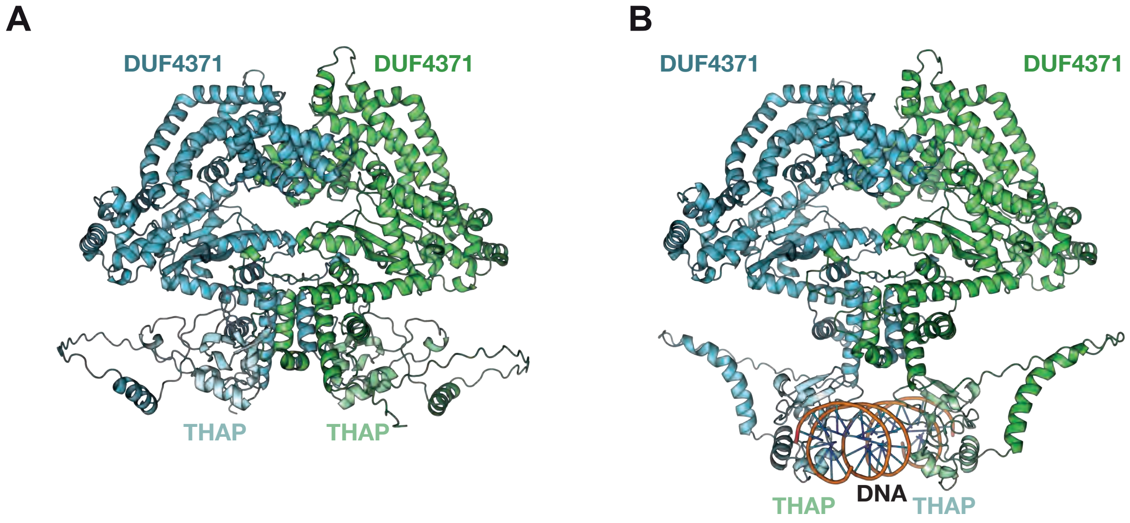

**Supplementary Figure S3. Efficient CRISPR-Cas9 mutagenesis of *thap12* paralogs in F0-CRISPant zebrafish. (A)** Schematic of *thap12a* and *thap12b* zebrafish paralogs showing conserved THAP and DUF4371 domains, along with CRISPR target sites (2 guide RNAs per ortholog). **(B-E)** High-resolution melting (HRM) analysis at different exons confirming successful genome editing in injected embryos for both *thap12a* **(B-C)** and *thap12b* **(D-E)** compared to sham-injected (5-mismatch gRNAs) and uninjected controls.

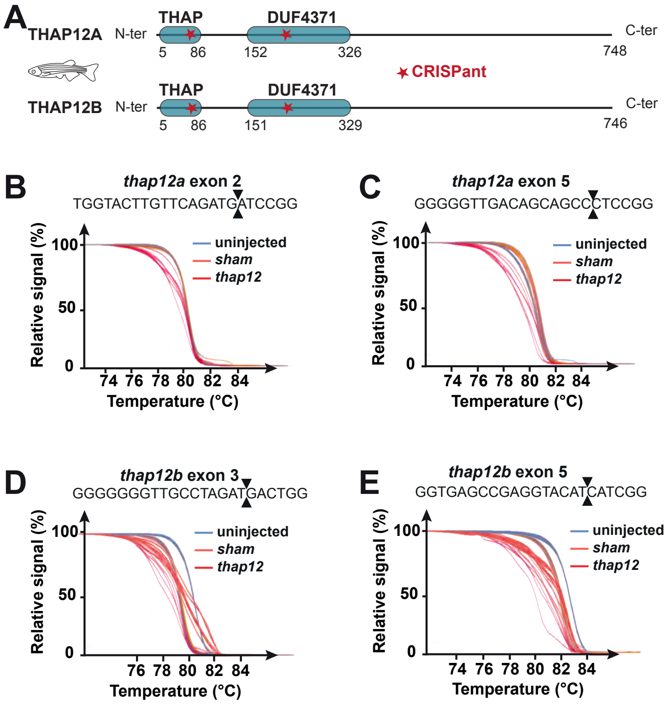

**Supplementary Figure S4. Functional connectivity alterations in *thap12*$^{CRISPant}$ larvae are robust and reproducible across experimental batches.** To assess the reproducibility of functional alterations caused by *thap12* knockdown, we first performed whole-brain two-photon calcium imaging in 16 *sham*-injected controls **(A)** and 16 *thap12*$^{CRISPant}$ mutants **(B)** at 5-6 days post-fertilization. For each larva, a maximum-intensity projection was generated from 21 imaging planes spanning the brain along the anterior-posterior axis, and raw fluorescence intensities were displayed on a consistent 16-bit scale to allow comparison across animals. To evaluate functional connectivity (FC) changes, we computed difference matrices representing the mean correlation changes between mutants and controls (ΔCorrelation) in two independent experimental batches. In batch 1 (**C**, $n$ = 9 per group) and batch 2 (**D**, $n$ = 7 per group), similar patterns of increased and decreased correlations were observed, suggesting a conserved FC signature in *thap12*$^{CRISPant}$ brains. To quantitatively assess inter-batch consistency, we plotted the FC difference values from batch 1 against those from batch 2 **(E)** and found a significant positive correlation (Pearson's $r$ = 0.353), indicating reproducibility of the observed network alterations. This empirical correlation (orange line in **F**) significantly exceeded the distribution of correlations generated by random shuffling of group labels across both batches (gray distribution; $P$ = 0.0197, 100,000 permutations with randomized experimental condition assignment), confirming that the observed FC changes are not due to chance or batch-specific effects.

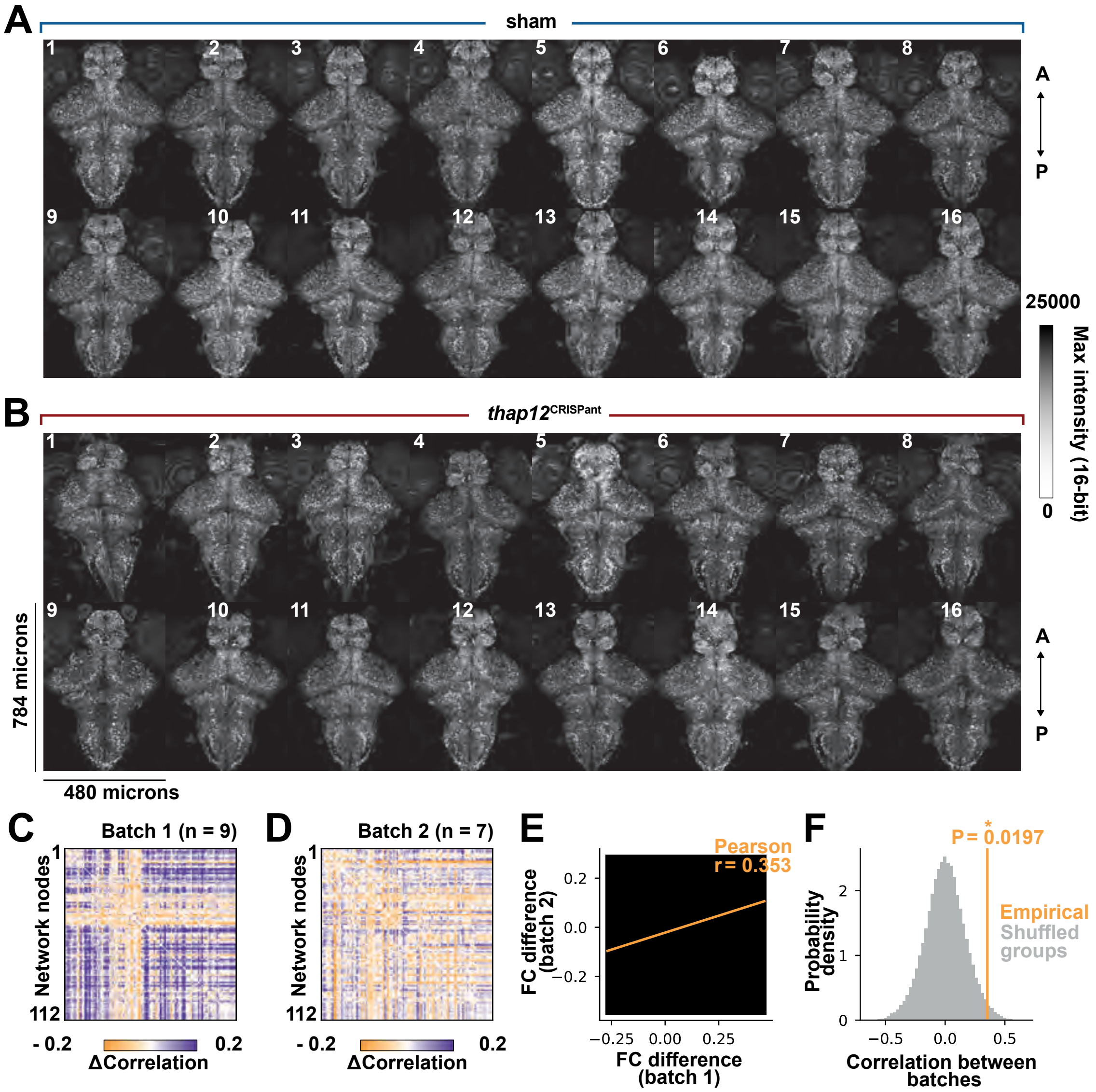

**Supplementary Figure S5 (next page). Stable CRISPR-CAS9 zebrafish mutants confirm that *thap12a*, but not *thap12b*, is essential for brain development and neuronal survival. (A-B)** Sanger sequencing confirms indel mutations in zebrafish *thap12a* (+20nt, **A**) and *thap12b* (del8nt, **B**) following CRISPR targeting within conserved domains. Both mutations introduce a premature stop codon at the end or within the N-terminal THAP domain (at position 57 in THAP12A and position 105 in THAP12B). **(C-F)**. Locomotor activity analysis under light (**C, E)** and dark **(D, F)** conditions shows significantly reduced swim distance in *thap12a*$^{-/-}$ larvae but not in *thap12b*$^{-/-}$ larvae at 4dpf. **(G)** Kaplan-Meier survival curves demonstrate early lethality at 15 dpf in *thap12a*$^{-/-}$ larvae but not in *thap12b*$^{-/-}$. **(H)** PTZ-induced seizure assays show increased hyperactivity in 4 dpf *thap12a*$^{-/-}$ but not *thap12b*$^{-/-}$ larvae (3 mM, 10 min). **(I)** Lateral images at 5 dpf illustrate reduced head size in *thap12a*$^{-/-}$ mutants. **(J-L)** Quantifications show unchanged body length **(J)**, but significantly reduced head length **(K)** and eye diameter **(L)** in *thap12a*$^{-/-}$ compared to wild-type siblings **(M)** Confocal images of 5 dpf *Tg(elavl3:GFP)* larvae reveal reduced brain volume in *thap12a*$^{-/-}$. **(N)** Quantification confirms a significant reduction in total brain area. **(O-P)** Caspase-3 immunostaining shows increased apoptosis in *thap12*$^{\text{CRISPant}}$ larvae compared to sham-injected controls. **(Q-R)** Phospho-Histone H3 (PH3) staining reveals reduced proliferative cell numbers in *thap12a*$^{-/-}$ larvae compared to +/+ siblings. **(S)** Additional quantification of caspase-3-positive cells confirms increased apoptosis in *thap12a*$^{-/-}$ larvae compared to +/+ siblings. *Statistical analyses in panels C-F used one-way ANOVA and in panels H, J-L, N, P, and R-S used unpaired two-tailed Student's t-test: *p < 0.05, **p < 0.01, ***p < 0.001, ****p < 0.0001; ns, not significant. (fb: forebrain; mb: midbrain; hb: hindbrain; bs:brainstem)*

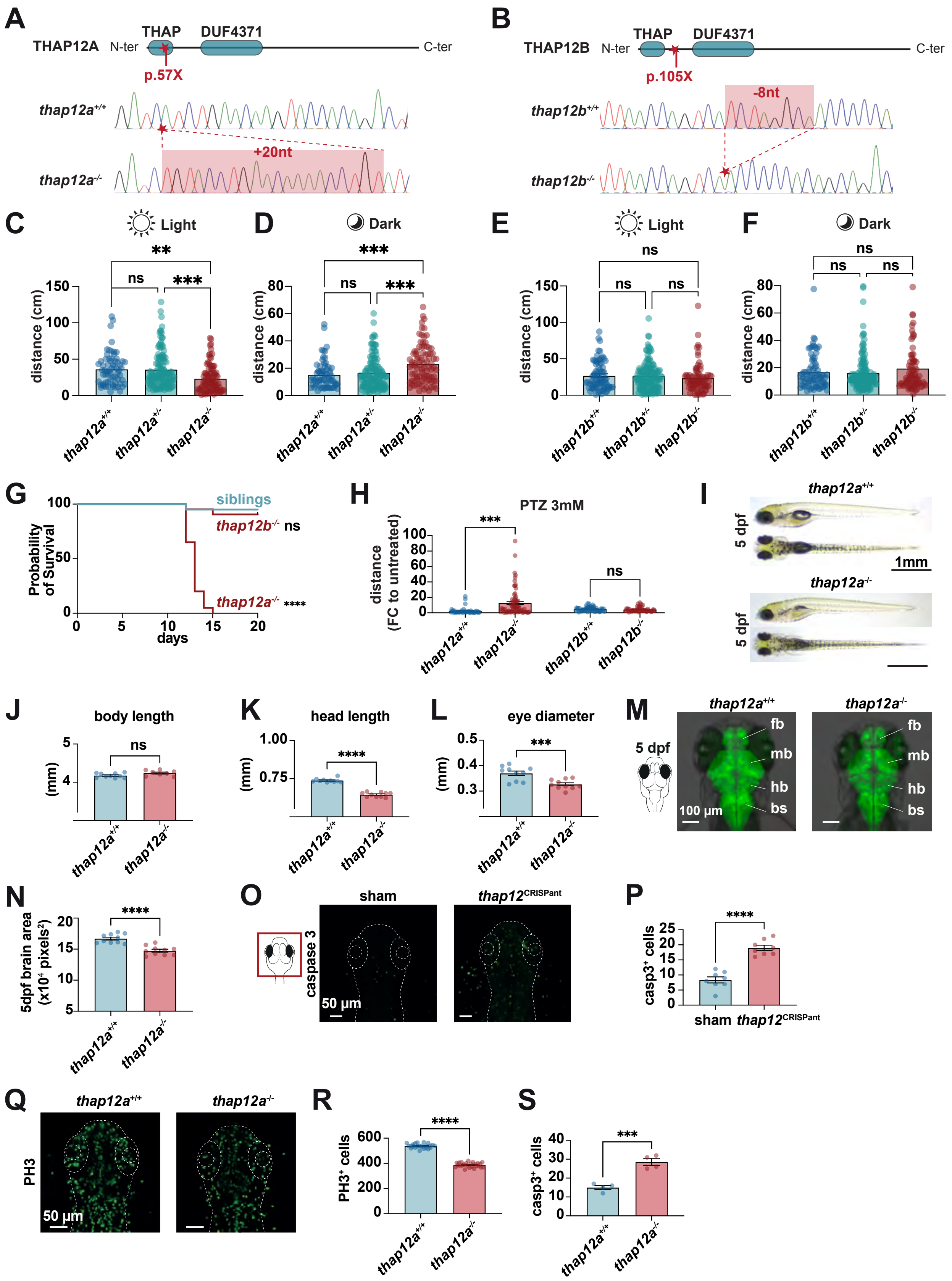

A
THAP12A
N-ter
THAP
DUF4371
C-ter
p.57X
thap12a+/+
thap12a-/-
+20nt
B
THAP12B
N-ter
THAP
DUF4371
C-ter
p.105X
-8nt
thap12b+/+
thap12b-/-
C
Light
D
Dark
E
Light
F
Dark
distance (cm)
G
siblings
thap12b-/- ns
thap12a-/- ****
Probability of Survival
days
H
PTZ 3mM
distance (FC to untreated)
I
thap12a+/+
thap12a-/-
5 dpf
1mm
J
body length
K
head length
L
eye diameter
(mm)
M
5 dpf
fb
mb
hb
bs
100 µm
N
5dpf brain area (x10⁴ pixels²)
O
sham
thap12CRISPant
caspase 3
50 µm
P
casp3+ cells
Q
PH3
50 µm
R
PH3+ cells
S
casp3+ cells

**Table S1. Differential expression in patient-derived fibroblasts.** RNA-seq analysis comparing primary fibroblasts from affected probands to healthy parents. The first sheet lists differentially expressed genes (DEGs), the second summarizes enriched biological pathways (Reactome GO).

**Table S2. Mouse genotyping data across development.** Raw genotyping counts and frequencies of *Thap12* knockout and compound heterozygous mice across developmental stages.

**Table S3. Differential expression in 4 dpf zebrafish brains.** RNA-seq analysis of microdissected brains from 4 dpf thap12a$^{-/-}$ zebrafish larvae versus wild-type siblings. The first tab lists differentially expressed genes (DEGs), and the second tab shows enriched KEGG pathways.

**Table S4. List of primers.**

**Video S1. Aberrant locomotion in *thap12*$^{CRISPants}$:** Representative video of *thap12*$^{CRISPants}$ larvae exhibiting erratic "hectic" high-speed movements during dark phases in behavioral assay.
